# Key factors to consider for *Candida auris* screening in healthcare settings: a systematic review

**DOI:** 10.1101/2024.11.06.24316836

**Authors:** Anders Skyrud Danielsen, Liz Ertzeid Ødeskaug, Ragnhild Raastad, Anne Kjerulf, Anne-Marie Andersen, Ragnhild Agathe Tornes, Jan P. W. Himmels, Ulf R. Dahle, Miriam Sare, Brian Kristensen, Hanne-Merete Eriksen-Volle, Mari Molvik

## Abstract

**Background:** *Candida auris* is an emerging fungal pathogen that is often multidrug-resistant. It can persist on skin and in hospital environments, leading to outbreaks and severe infections for patients at risk. Several countries and institutions are working on establishing guidelines and recommendations for prevention. This review aims to assess the evidence on factors associated with *C. auris* colonisation or infection, the duration of such colonisation, possible colonisation sites, and the risk of secondary cases to inform screening recommendations.

**Methods:** We systematically searched five databases for primary studies and systematic reviews of our four outcomes. We excluded studies on treatment, management, laboratory methods, drug resistance, and environmental screening. From each paper, we extracted relevant data and summarised them in tables. Main findings were described narratively.

**Findings:** We selected 117 studies for inclusion. Most of the studies were observational studies. Without taking the method of testing into account, the duration of *C. auris* colonisation varied, with up to and beyond a year being common. The predominant sites of colonisation were the axillae and groin, with the nares and rectum being less common sites. The risk of secondary cases saw considerable variation across the studies, and the secondary cases primarily involved patients and not health care workers. Critical care settings, invasive medical devices, recent antimicrobial use, and comorbidities were often associated with *C. auris* colonisation and infection.

**Conclusion:** Our review highlights that, despite relevant findings on factors influencing *C. auris* colonisation and infection, substantial gaps remain in the evidence supporting screening practices. Most studies were conducted reactively, in outbreak settings, and lack systematic protocols. Given these limitations, screening guidelines are likely to be more successful if grounded in medical theory and yeast microbiology rather than relying solely on current studies. Rigorous, well-designed research is urgently needed to inform future *C. auris* screening and control efforts.

## Background

*Candida auris* is a fungal pathogen that rapidly emerged globally. While initially isolated from the external ear canal of a Japanese patient in 2009 and retrospectively identified in South Korean blood samples dating back to 1996, the first large hospital outbreak triggering worldwide concern occurred in India and was described in 2013 (2–5). In the 2019 Antibiotic Resistance Threat Report by the U.S Centers for Disease Control and Prevention (CDC), *C. auris* was listed as an urgent threat and in 2022 the World Health Organization (WHO) classified it as a critical priority pathogen (6;7). The urgency communicated from these institutions stems from the rapid emergence of *C. auris*, its resistance to antifungal treatments, and its ability to cause outbreaks and severe, often fatal, infections in hospitalised patients, particularly those with underlying conditions(7).

*C. auris* can colonise and persist on the skin for longer periods and exhibits extended environmental survival on inanimate surfaces within healthcare settings. These characteristics contribute to transmission events and protracted outbreaks in healthcare settings, especially in intensive care units (ICU) (2;8). Furthermore, *C. auris* is capable of biofilm formation and can resist routine cleaning (9–11). Four major clades – South Asian (I), East Asian (II), South African (III) and South American (IV) – have accounted for the global spread of the pathogen (2). Clade I is the most prevalent. Genetic studies using whole genome sequencing (WGS) suggest a simultaneous emergence of these clades, which may have played a part in causing the rapid global spread. Recently one more clade; Iranian (V) has been identified, and a sixth clade is under investigation in Singapore (VI) (2;12). Variations in drug resistance and virulence, the ability to cause invasive disease, have been observed among different *C. auris* clades (2).

Although *C. auris* has been reported in over 60 countries across six continents since its discovery (9;13), case numbers likely underestimate the true burden as the fungus has proven notoriously difficult to culture (14). The first identified case of *C. auris* in Europe, belonging to clade I, was imported from India in 2007 and retrospectively identified (15). Since then, several European countries have witnessed a significant increase in cases, causing outbreaks, especially in ICU (2). Between 2013 and 2021, 15 EU/EEA countries identified a total of 1812 cases (11). The first European outbreak occurred in 2015-2016 in an ICU in the United Kingdom with a total of 70 cases (16). Between 2019 and 2021, five European countries (Denmark, France, Germany, Greece and Italy) reported a total of 14 *C. auris* outbreaks, encompassing 327 cases (17).

Several national and local institutions are moving to either recommend or mandate screening against *C. auris* in their prevention effort, e.g., in guideline documents. This highlights the need to assess the evidence on some key factors to consider when developing screening strategies.

These key factors are the associated factors of colonisation/infection, duration of colonisation, possible colonisation sites and risk of secondary cases. This systematic review aimed to comprehensively search for and narratively summarise the available literature on these key factors, including all relevant studies published to date.

## Methods

### Search and study selection

We systematically searched five databases on 2 February 2024: Embase, Ovid Medline, Cochrane Database of Systematic Reviews/ Cochrane Central Register of Controlled Trials, Web of Science, and Epistemonikos. The searches were complemented with hand-searches for grey literature and existing guidelines. The searches were performed by a specialist librarian, after internal peer review by another librarian. Search terms for *C. auris* combined with synonyms with appropriate truncations and abbreviations were used for searching title, abstract, author keywords, and controlled vocabulary. The search strategy was tailored to each database’s search interface. The search strategy can be found in Appendix 1. Deduplication was performed in EndNote 20 (18).

Three researchers piloted the inclusion and exclusion criteria on an initial sample of 50 abstracts to identify relevant inclusion criteria (Table 1). Subsequently, two researchers screened the remaining studies. We used EPPI-Reviewer 6 for screening (19). Disagreements or uncertainties were addressed through discussion with a third researcher. Two researchers performed single full-text screening. Inclusion conflicts were resolved through discussion between the researchers.

**Table 1.** Inclusion and exclusion criteria.

| Inclusion criteria | Exclusion criteria |
| --- | --- |
| - Individuals tested/screened for *C. auris*<br>- Studies reporting duration of *C. auris* colonisation<br>- Studies reporting number of secondary cases in relation to a positive *C. auris* case<br>- Studies reporting factors associated with *C. auris* infection or colonisation<br>- Studies with reported screening results by body site<br>- Systematic reviews with systematic literature search and pre-defined inclusion criteria<br>- RCT and observational studies with total participants $\leq 5$<br>- No limitations on year of publication<br>- No filter on country/context for reviews and trials<br>- Studies in English, Norwegian, Swedish, or Danish | - Studies on treatment of *C. auris*<br>- Studies on management of *C. auris* cases<br>- Studies on laboratory methods (including sampling methods) for detecting *C. auris*<br>- Studies investigating drug resistance in *C. auris*<br>- Studies related to the genetics or cell biology of *C. auris*<br>- Studies exclusively focused on environmental screening or environmental sampling during outbreaks<br>- Studies investigating preventive measures, including screening, against postoperative wound infections<br>- Cross-sectional studies featuring aggregated data (considered for studies on colonisation sites)<br>- Case reports with $n \leq 5$ *C. auris* positive cases<br>- Letters to the editor, abstracts/posters, non-peer-reviewed studies, correspondences, short communications, and comments<br>- Studies where outcomes are not specifically reported for *C. auris* |

### Data extraction and analysis

Two researchers extracted relevant data on three of the main outcomes: duration of colonisation, risk of secondary cases, and colonisation sites. One researcher extracted data regarding associated factors. For all studies we extracted information on the country of study, study period, setting, participants, follow-up period, and reported outcomes. Additionally, depending on the specific outcome of interest, we also extracted relevant data and statistics (e.g., odds ratio).

For each outcome we created tables summarising the relevant studies. Each table lists the studies reporting on the specific outcomes, along with the variables relevant for the outcome. There was a limited number of studies featuring control groups that investigated associated factors or participant characteristics, and only these were included in the interpreting and assessing factors associated with infection or colonisation. We narratively summarised each of the systematic reviews separately, and then findings for each outcome from the primary studies, where similar studies or studies reporting on comparable aspects to each outcome were grouped together for easier comparison. To assist in the synthesis and discussion we employed a set of definitions (Table 2). Neither a meta-analysis or a critical appraisal/quality assessment of the studies was performed due to the qualitative heterogeneity in the primary studies – both in terms of design and context. However, we highlighted methodological shortcomings throughout the text to provide context for our findings.

**Table 2.**
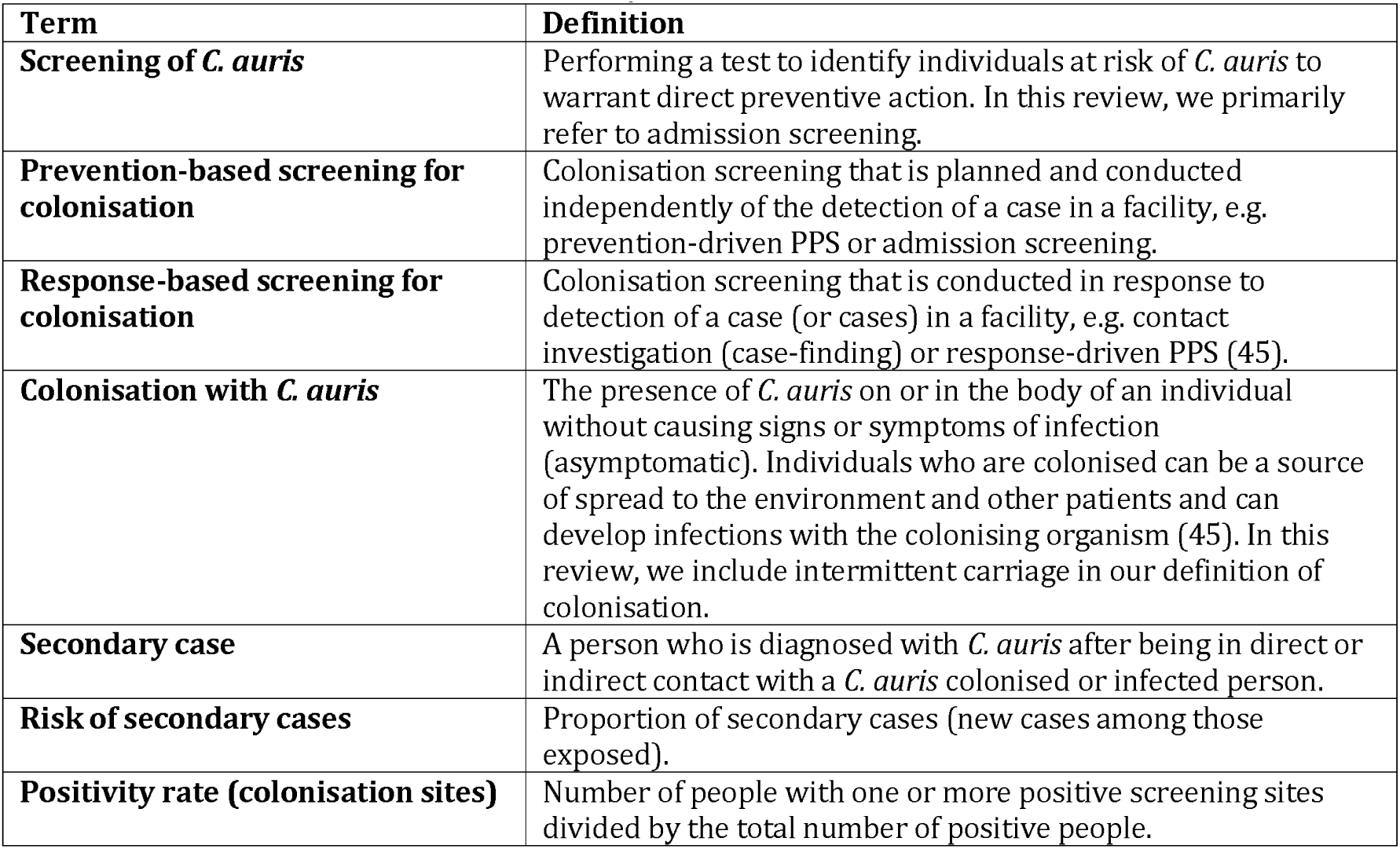
Definitions used in the narrative synthesis and discussion.

## Findings

### Selected studies

We identified 2,371 unique references after deduplication (Figure 1). Title and abstract screening identified 291 relevant studies. On full-text screening we included 117 studies. Studies were conducted in Asia (n=40), North America (n=29), Europe (n=21), South America (n=12), Africa (n=8), and Oceania (n=1). Six studies encompassed data from multiple countries.

**Figure 1:**
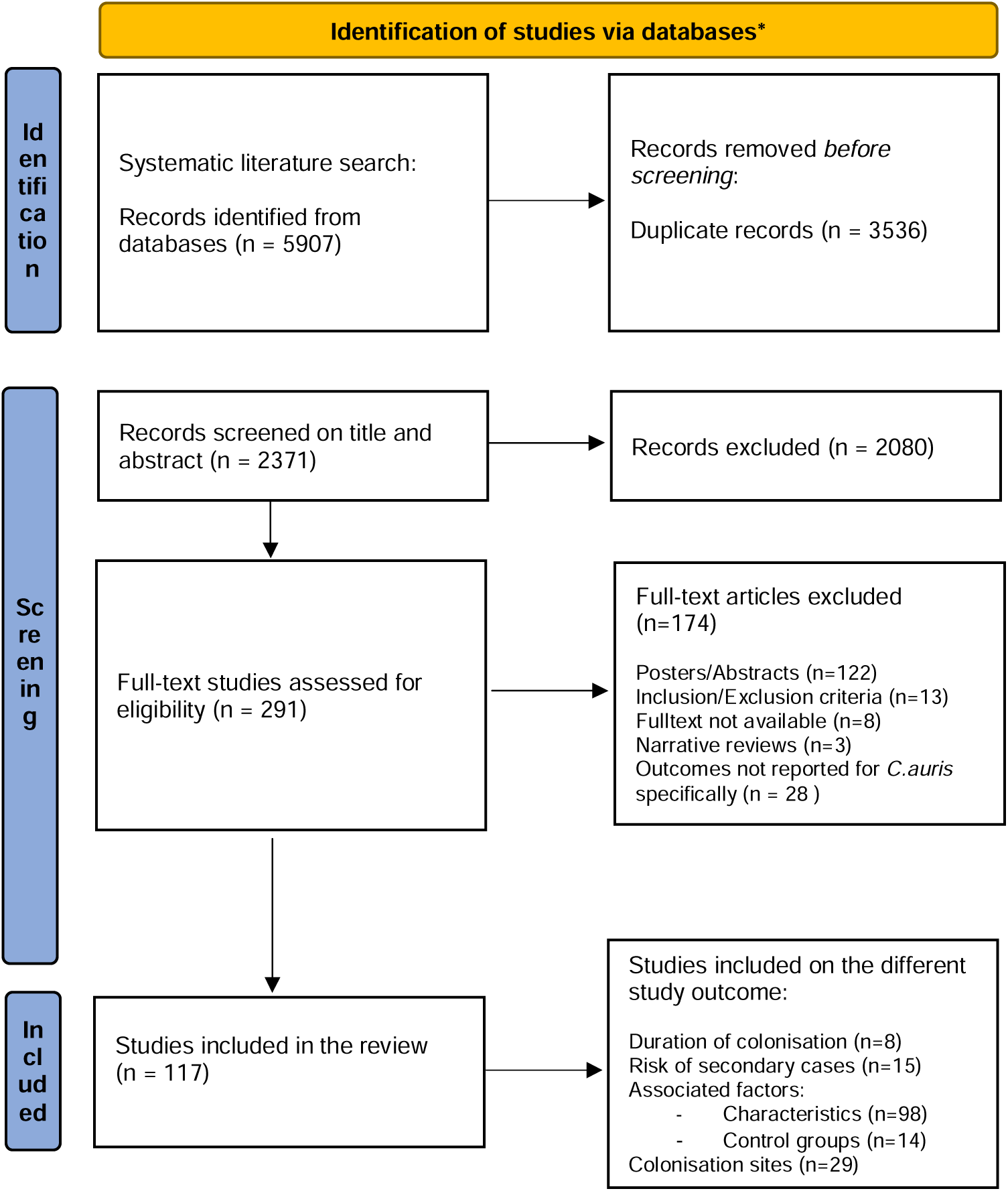
Flow diagram of search strategy and study inclusion. Adapted from (1). *Databases include Ovid Medline® and Epub Ahead of Print, Embase, Cochrane Database of Systematic Reviews, Cochrane Central Register of Controlled Trials, Web of Science and Epistemonikos.

Publication dates ranged from 2016 to 2024. We included 114 observational studies and three systematic reviews. Reported outcomes of interest were duration of colonisation (n=8), risk of secondary cases (n=15), colonisation sites (n=29), and associated factors/participant characteristics (n=112). Only a limited number of studies that investigated associated factors or participant characteristics featured control groups (n=14). In general, the included studies exhibited considerable variation in study populations and sample sizes. The body of literature on *C. auris* appear to primarily consist of single-centre studies with small sample sizes and descriptive statistics. These are often outbreak reports, summaries of experiences with the clinical management of defined clusters, or studies summarising data from laboratory findings over a specific period. An overview of the included studies is provided in Table 3.

**Table 3.** Overview of included studies.

| Author (Year) | Title | Country | Outcome reported |
| --- | --- | --- | --- |
| **Systematic reviews** |  |  |  |
| Osei Sekyere J (2018) (8) | Candida auris: A systematic review and meta-analysis of current updates on an emerging multidrug-resistant pathogen | Several | Associated factors (characteristics) |
| Vaseghi N (2022) (20) | Global prevalence and subgroup analyses of coronavirus disease (COVID-19) associated Candida auris infections (CACA): A systematic review and meta-analysis | Several | Associated factors (characteristics) |
| Vinayagamoorthy K (2022) (21) | Prevalence, risk factors, treatment and outcome of multidrug resistance Candida auris infections in Coronavirus disease (COVID-19) patients: A systematic review | Most cases from USA, Mexico, India | Associated factors (characteristics) |
| **Observational studies** |  |  |  |
| Adam RD (2019) (48) | Analysis of Candida auris fungemia at a single facility in Kenya | Kenya | Associated factors (control group) |
| Adams E (2018) (25) | Candida auris in Healthcare Facilities, New York, USA, 2013-2017 | USA | Duration of colonisation, risk of secondary cases, colonisation site, associated factors (characteristics) |
| Ahmad S (2020) (83) | Candida auris in various hospitals across Kuwait and their susceptibility and molecular basis of resistance to antifungal drugs | Kuwait | Associated factors (characteristics) |
| Al Maani A (2019) (84) | Ongoing Challenges with Healthcare-Associated Candida auris Outbreaks in Oman | Oman | Associated factors (characteristics) |
| Alfouzan W (2020) (85) | Molecular Epidemiology of Candida Auris Outbreak in a Major Secondary-Care Hospital in Kuwait | Kuwait | Associated factors (characteristics) |
| Allaw F (2022) (49) | COVID-19 and C. auris: A Case-Control Study from a Tertiary Care Center in Lebanon | Lebanon | Associated factors (control group) |
| Allaw F (2021) (86) | First Candida auris Outbreak during a COVID-19 Pandemic in a Tertiary-Care Center in Lebanon | Lebanon | Associated factors (characteristics) |
| Almaghrabi RS (2020) (87) | Molecular characterisation and clinical outcomes of Candida auris infection: Single-centre experience in Saudi Arabia | Saudi Arabia | Associated factors (characteristics) |
| Al-Rashdi A (2021) (88) | Characteristics, Risk Factors, and Survival Analysis of Candida auris Cases: Results of One-Year National Surveillance Data from Oman | Oman | Associated factors (characteristics) |
| Alshahrani FS (2023) (79) | Description of Candida auris Occurrence in a Tertiary Health Institution in Riyadh, Saudi Arabia | Saudi Arabia | Colonisation site, associated factors (characteristics) |
| Alshamrani MM (2021) (35) | Management of Candida auris outbreak in a tertiary-care setting in Saudi Arabia | Saudi Arabia | Risk of secondary cases, associated factors (characteristics) |
| Alvarado-Socarras JL (2021) (89) | A Cluster of Neonatal Infections Caused by Candida auris at a Large Referral Center in Colombia | Colombia | Associated factors (characteristics) |
| Alvarez-Moreno CA (2023) (50) | The Mortality Attributable to Candidemia in C. auris Is Higher than That in Other Candida Species: Myth or Reality? | Colombia | Associated factors (control group) |
| Amer HA (2023) (90) | Characteristics and Mitigation Measures of Candida auris Infection: Descriptive Analysis from a Quaternary Care Hospital in Saudi Arabia, 2021-2022 | Saudi Arabia | Associated factors (characteristics) |
| Arauz AB (2018) (91) | Isolation of Candida auris from 9 patients in Central America: Importance of accurate diagnosis and susceptibility testing | Panama | Associated factors (characteristics) |
| Arenas SP (2023) (26) | Persistent colonisation of Candida auris among inpatients rescreened as part of a weekly surveillance program | USA | Duration of colonisation, associated factors (characteristics) |
| Arensman K (2020) (92) | Clinical Outcomes of Patients Treated for Candida auris Infections in a Multisite Health System, Illinois, USA | USA | Associated factors (characteristics) |
| Armstrong PA (2019) (70) | Hospital-Associated Multicenter Outbreak of Emerging Fungus Candida auris, Colombia, 2016 | Colombia | Colonisation site, associated factors (characteristics) |
| Asadzadeh M (2022) (93) | Molecular characterisation of Candida auris isolates from immunocompromised patients in a tertiary-care hospital in Kuwait reveals a novel mutation in FKS1 conferring reduced susceptibility to echinocandins | Kuwait | Associated factors (characteristics) |
| Barantsevich NE (2019) (94) | Emergence of Candida auris in Russia | Russia | Associated factors (characteristics) |
| Barantsevich NE (2020) (95) | Candida auris Bloodstream Infections in Russia | Russia | Associated factors (characteristics) |
| Benedict K (2023) (96) | Candida auris-Associated Hospitalizations, United States, 2017-2022 | USA | Associated factors (characteristics) |
| Bergeron G (2021) (22) | Candida auris Colonisation After Discharge to a Community Setting: New York City, 2017-2019 | USA | Duration of colonisation, associated factors (characteristics) |
| Berrio I (2021) (97) | Bloodstream Infections with Candida auris Among Children in Colombia: Clinical Characteristics and Outcomes of 34 Cases | Colombia | Associated factors (characteristics) |
| Bing J (2024) (98) | Candida auris-associated hospitalizations and outbreaks, China, 2018-2023 | China | Associated factors (characteristics) |
| Biran R (2023) (99) | Nationwide Outbreak of Candida auris Infections Driven by COVID-19 Hospitalizations, Israel, 2021-2022 | Israel | Associated factors (characteristics) |
| Biswal M (2017) (38) | Controlling a possible outbreak of Candida auris infection: lessons learnt from multiple interventions | India | Risk of secondary cases, colonisation site |
| Briano F (2022) (100) | Candida auris Candidemia in Critically Ill, Colonised Patients: Cumulative Incidence and Risk Factors | Italy | Colonisation site, associated factors (characteristics) |
| Caceres DH (2020) (51) | Case-Case Comparison of Candida auris Versus Other Candida Species Bloodstream Infections: Results of an Outbreak Investigation in Colombia | Colombia | Associated factors (control group) |
| Calvo B (2016) (101) | First report of Candida auris in America: Clinical and microbiological aspects of 18 episodes of candidemia | Venezuela | Associated factors (characteristics) |
| Chakrabarti A (2020) (102) | Characteristics, outcome and risk factors for mortality of paediatric patients with ICU-acquired candidemia in India: A multicentre prospective study | India | Associated factors (characteristics) |
| Chandramati J (2020) (103) | Neonatal Candida auris infection: Management and prevention strategies - A single centre experience | India | Associated factors (characteristics) |
| Chibabhai V (2022) (104) | Incidence of candidemia and prevalence of azole-resistant candidemia at a tertiary South African hospital - A retrospective laboratory analysis 2016-2020 | South Africa | Associated factors (characteristics) |
| Chow NA (2018) (105) | Multiple introductions and subsequent transmission of multidrug-resistant Candida auris in the USA: a molecular epidemiological survey | USA | Colonisation site, associated factors (characteristics) |
| Chowdhary A (2020) (106) | Multidrug-Resistant Candida auris Infections in Critically Ill Coronavirus Disease Patients, India, April-July 2020 | India | Associated factors (characteristics) |
| Corcione S (2022) (107) | First Cases of Candida auris in a Referral Intensive Care Unit in Piedmont Region, Italy | Italy | Colonisation site, associated factors (characteristics) |
| de Melo CC (2023) (71) | Colonised patients by Candida auris: Third and largest outbreak in Brazil and impact of biofilm formation | Brazil | Colonisation site, associated factors (characteristics) |
| de St Maurice A (2023) (108) | Clinical, microbiological, and genomic characteristics of clade-III Candida auris colonisation and infection in southern California, 2019-2022 | USA | Associated factors (characteristics) |
| Di Pilato V (2021) (109) | Molecular Epidemiological Investigation of a Nosocomial Cluster of C. auris: Evidence of Recent Emergence in Italy and Ease of Transmission during the COVID-19 Pandemic | Italy | Risk of secondary cases |
| Eckbo EJ (2021) (36) | First reported outbreak of the emerging pathogen Candida auris in Canada | Canada | Risk of secondary cases, associated factors (characteristics) |
| Escandon P (2018) (110) | Notes from the Field: Surveillance for Candida auris - Colombia, September 2016-May 2017 | Colombia | Colonisation site, associated factors (characteristics) |
| Escandon P (2022) (111) | Laboratory-based surveillance of Candida auris in Colombia, 2016-2020 | Colombia | Associated factors (characteristics) |
| Escandon P (2019) (34) | Molecular Epidemiology of Candida auris in Colombia Reveals a Highly Related, Countrywide Colonisation with Regional Patterns in Amphotericin B Resistance | Colombia | Risk of secondary cases, associated factors (characteristics) |
| Eyre DW (2018) (16) | A Candida auris Outbreak and Its Control in an Intensive Care Setting | UK | Duration of colonisation, risk of secondary cases, colonisation site, associated factors (characteristics) |
| Farooqi JQ (2020) (52) | Outbreak investigation of Candida auris at a tertiary care hospital in Karachi, Pakistan | Pakistan | Associated factors (control group) |
| Garcia-Bustos V (2020) (112) | A clinical predictive model of candidaemia by Candida auris in previously colonised critically ill patients | Spain | Colonisation site, associated factors (characteristics) |
| Garcia-Jeldes HF (2020) (113) | Prevalence of Candida auris in Canadian acute care hospitals among at-risk patients, 2018 | Canada | Associated factors (characteristics) |
| Gómez CF (2021) (114) | Analysis of Candida auris candidemia cases in an Intensive Care Unit of a tertiary hospital | Spain | Associated factors (characteristics) |
| Goulart MA (2023) (72) | Identification and infection control response to Candida auris at an academic level I trauma center | USA | Colonisation site, associated factors (characteristics) |
| Govender NP (2018) (115) | Candida auris in South Africa, 2012-2016 | South Africa | Associated factors (characteristics) |
| Hamprecht A (2019) (65) | Candida auris in Germany and Previous Exposure to Foreign Healthcare | Germany | Associated factors (characteristics) |
| Hanson BM (2021) (116) | Candida auris Invasive Infections during a COVID-19 Case Surge | USA | Associated factors (characteristics) |
| Jung J (2020) (24) | Candida auris colonisation or infection of the ear: A single-center study in South Korea from 2016 to 2018 | South Korea | Duration of colonisation, associated factors (characteristics) |
| Kaki R (2023) (117) | Risk factors and mortality of the newly emerging Candida auris in a university hospital in Saudi Arabia | Saudi Arabia | Associated factors (characteristics) |
| Kekana D (2023) (118) | Candida auris Clinical Isolates Associated with Outbreak in Neonatal Unit of Tertiary Academic Hospital, South Africa | South Africa | Associated factors (characteristics) |
| Khan Z (2018) (119) | Invasive Candida auris infections in Kuwait hospitals: epidemiology, antifungal treatment and outcome | Kuwait | Associated factors (characteristics) |
| Koleri J (2023) (120) | Candida auris Blood stream infection - a descriptive study from Qatar | Qatar | Associated factors (characteristics) |
| Lee EH (2024) (39) | Intrahospital transmission and infection control of Candida auris originating from a severely infected COVID-19 patient transferred abroad | South Korea | Risk of secondary cases, colonisation site, associated factors (characteristics) |
| Leonhard SE (2024) (63) | Proposal for a screening protocol for Candida auris colonisation | Netherlands | Colonisation site, associated factors (characteristics) |
| Lockhart SR (2017) (121) | Simultaneous Emergence of Multidrug-Resistant Candida auris on 3 Continents Confirmed by Whole-Genome Sequencing and Epidemiological Analyses | Pakistan, India, Venezuela, South Africa | Associated factors (characteristics) |
| Magnasco L (2021) (122) | Spread of Carbapenem-Resistant Gram-Negatives and Candida auris during the COVID-19 Pandemic in Critically Ill Patients: One Step Back in Antimicrobial Stewardship? | Italy | Associated factors (characteristics) |
| Magnasco L (2023) (123) | Frequency of Detection of Candida auris Colonisation Outside a Highly Endemic Setting: What Is the Optimal Strategy for Screening of Carriage? | Italy | Associated factors (characteristics) |
| Magobo R (2020) (124) | Multilocus sequence typing of azole-resistant Candida auris strains, South Africa | South Africa | Associated factors (characteristics) |
| McDougal AN (2023) (42) | A cluster investigation of Candida auris among hospitalized incarcerated patients | USA | Risk of secondary cases, colonisation site, associated factors (characteristics) |
| McPherson TD (2020) (73) | Notes from the Field: Candida auris and Carbapenemase-Producing Organism Prevalence in a Pediatric Hospital Providing Long-Term Transitional Care - Chicago, Illinois, 2019 | USA | Colonisation site, Risk factors (characteristics) |
| Mohsin J (2020) (125) | A Cluster of Candida auris Blood Stream Infections in a Tertiary Care Hospital in Oman from 2016 to 2019 | Oman | Associated factors (characteristics) |
| Moin S (2021) (53) | C. auris and non-C. auris candidemia in hospitalized adult and pediatric COVID-19 patients; single center data from Pakistan | Pakistan | Associated factors (control group) |
| Morales-Lopez SE (2017) (126) | Invasive Infections with Multidrug-Resistant Yeast Candida auris, Colombia | Colombia | Associated factors (characteristics) |
| Mulet Bayona JV (2023) (127) | Candida auris from colonisation to candidemia: A four-year study | Spain | Associated factors (characteristics) |
| Mulet Bayona JV (2020) (128) | Characteristics and Management of Candidaemia Episodes in an Established Candida auris Outbreak | Spain | Colonisation site, associated factors (characteristics) |
| Munshi A (2024) (129) | Risk factors, antifungal susceptibility, complications, and outcome of Candida auris bloodstream infection in a tertiary care center in the western region of Saudi Arabia | Saudi Arabia | Associated factors (characteristics) |
| Nobrega de Almeida J (2021) (130) | Axillary Digital Thermometers uplifted a multidrug-susceptible Candida auris outbreak among COVID-19 patients in Brazil | Brazil | Associated factors (characteristics) |
| Ortiz-Roa C (2023) (131) | Mortality Caused by Candida auris Bloodstream Infections in Comparison with Other Candida Species, a Multicentre Retrospective Cohort | Colombia | Associated factors (characteristics) |
| Pacilli M (2020) (27) | Regional Emergence of Candida auris in Chicago and Lessons Learned From Intensive Follow-up at 1 Ventilator-Capable Skilled Nursing Facility | USA | Duration of colonisation, transmission, associated factors (characteristics) |
| Pandya N (2021) (132) | International Multicentre Study of Candida auris Infections | India, Oman, Turkey, USA, Pakistan | Associated factors (characteristics) |
| Parak A (2022) (54) | Clinical and laboratory features of patients with Candida auris cultures, compared to other Candida, at a South African Hospital | South Africa | Associated factors (control group) |
| Park JY (2019) (133) | Management of Patients with Candida auris Fungemia at Community Hospital, Brooklyn, New York, USA, 2016-2018<sup>1</sup> | USA | Associated factors (characteristics) |
| Peng Y (2024) (134) | First report of Candida auris in Guangdong, China: clinical and microbiological characteristics of 7 episodes of candidemia | China | Associated factors (characteristics) |
| Piatti G (2022) (78) | Colonisation by Candida auris in critically ill patients: role of cutaneous and rectal localization during an outbreak | Italy | Colonisation site, associated factors (characteristics) |
| Prestel C (2021) (135) | Candida auris Outbreak in a COVID-19 Specialty Care Unit - Florida, July-August 2020 | USA | Associated factors (characteristics) |
| Proctor DM (2021) (77) | Integrated genomic, epidemiologic investigation of Candida auris skin colonisation in a skilled nursing facility | USA | Colonisation site, associated factors (characteristics) |
| Rossow J (2021) (31) | Factors Associated with Candida auris Colonisation and Transmission in Skilled Nursing Facilities With Ventilator Units, New York, 2016-2018 | USA | Risk of secondary cases, associated factors (control group) |
| Rowlands J (2023) (75) | Candida auris admission screening pilot in select units of New York City health care facilities, 2017-2019 | USA | Colonisation site, associated factors (characteristics) |
| Ruiz-Gaitan A (2019) (55) | Detection and treatment of Candida auris in an outbreak situation: risk factors for developing colonisation and candidemia by this new species in critically ill patients | Spain | Associated factors (control group) |
| Ruiz-Gaitan A (2018) (41) | An outbreak due to Candida auris with prolonged colonisation and candidaemia in a tertiary care European hospital | Spain | Colonisation site, secondary transmission, associated factors (characteristics) |
| Sathyapalan DT (2021) (136) | Evaluating the measures taken to contain a Candida auris outbreak in a tertiary care hospital in South India: an outbreak investigational study | India | Associated factors (characteristics) |
| Sayeed MA (2019) (137) | Clinical spectrum and factors impacting outcome of Candida auris: a single center study from Pakistan | Pakistan | Associated factors (characteristics) |
| Sayeed MA (2020) (56) | Comparison of risk factors and outcomes of Candida auris candidemia with non-Candida auris candidemia: A retrospective study from Pakistan | Pakistan | Associated factors (control group) |
| Schelenz S (2016) (40) | First hospital outbreak of the globally emerging Candida auris in a European hospital | UK | Risk of secondary cases, colonisation site |
| Sharp A (2021) (138) | Screening for Candida auris in patients admitted to eight intensive care units in England, 2017 to 2018 | UK | Colonisation site |
| Shastri PS (2020) (57) | Candida auris candidaemia in an intensive care unit - Prospective observational study to evaluate epidemiology, risk factors, and outcome | India | Associated factors (control group) |
| Shaukat A (2021) (139) | Experience of treating Candida auris cases at a general hospital in the state of Qatar | Qatar | Associated factors (characteristics) |
| Simon SP (2023) (58) | Comparative Outcomes of Candida auris Bloodstream Infections: A Multicenter Retrospective Case-Control Study | USA | Associated factors (control group) |
| Southwick K (2022) (74) | A description of the first Candida auris-colonised individuals in New York State, 2016-2017 | USA | Colonisation site, associated factors (characteristics) |
| Stanciu (2023) (140) | First report of Candida auris in Romania: clinical and molecular aspects | Romania | Associated factors (characteristics) |
| Sticchi C (2023) (141) | Increasing Number of Cases Due to Candida auris in North Italy, July 2019-December 2022 | Italy | Associated factors (characteristics) |
| Taori SK (2019) (59) | Candida auris outbreak: Mortality, interventions and cost of sustaining control | UK | Associated factors (control group) |
| Taori SK (2022) (142) | First experience of implementing Candida auris real-time PCR for surveillance in the UK: detection of multiple introductions with two international clades and improved patient outcomes | UK | Associated factors (characteristics) |
| Thomsen J (2023) (143) | Emergence of highly resistant Candida auris in the United Arab Emirates: a retrospective analysis of evolving national trends | United Arab Emirates | Associated factors (characteristics) |
| Tian S (2018) (144) | First cases and risk factors of super yeast Candida auris infection or colonisation from Shenyang, China | China | Associated factors (characteristics) |
| Townsend JO (2021) (33) | Identification of Candida auris in a foreign repatriated patient to Ontario, Canada and infection control strategies to prevent transmission | Canada | Secondary transmission, colonisation site |
| Tsay S (2017) (145) | Notes from the Field: Ongoing Transmission of Candida auris in Health Care Facilities - United States, June 2016-May 2017 | USA | Colonisation site, associated factors (characteristics) |
| Turbett IS (2022) (66) | Evaluation of Candida auris acquisition in US international travellers using a culture-based screening protocol | Africa, Asia, USA, Europe | Colonisation site, associated factors (characteristics) |
| Umamaheshwari S (2021) (146) | Clinical profile, antifungal susceptibility, and molecular characterization of Candida auris isolated from patients in a South Indian surgical ICU | India | Risk factors (characteristics) |
| Vallabhaneni S (2016) (23) | Investigation of the First Seven Reported Cases of Candida auris, a Globally Emerging Invasive, Multidrug-Resistant Fungus-United States, May 2013-August 2016 | USA | Duration of colonisation, colonisation site, associated factors (characteristics) |
| van Schalkwyk E (2019) (60) | Epidemiologic Shift in Candidemia Driven by Candida auris, South Africa, 2016-2017 | South Africa | Associated factors (control group) |
| Villanueva-Lozano H (2021) (147) | Outbreak of Candida auris infection in a COVID-19 hospital in Mexico | Mexico | Associated factors (characteristics) |
| Vu CA (2022) (148) | Challenges and opportunities in stewardship among solid organ transplant recipients with Candida auris bloodstream infections | USA | Associated factors (characteristics) |
| Walits E (2023) (149) | Outcome of Candida auris contact investigations conducted in a 6 month period at a New York City hospital | USA | Associated factors (characteristics) |
| Waters A (2023) (150) | Investigation of a Candida auris outbreak in a skilled nursing facility - Virginia, United States, October 2020-June 2021 | USA | Associated factors (characteristics) |
| Worth LJ (2020) (37) | Candida auris in an Australian health care facility: importance of screening high risk patients | Australia | Secondary transmission, associated factors (characteristics) |
| Yadav A (2021) (28) | Colonisation and Transmission Dynamics of Candida auris among Chronic Respiratory Diseases Patients Hospitalised in a Chest Hospital, Delhi, India: A Comparative Analysis of Whole Genome Sequencing and Microsatellite Typing | India | Duration of colonisation, colonisation site, associated factors (characteristics) |
| Zerrouki H (2022) (151) | Emergence of Candida auris in intensive care units in Algeria | Algeria | Associated factors (characteristics) |
| Zhu Y (2020) (76) | Laboratory Analysis of an Outbreak of Candida auris in New York from 2016 to 2018: Impact and Lessons Learned | USA | Colonisation site, associated factors (characteristics) |

### Systematic reviews

We identified three systematic reviews (8;20;21). Sekyere authored a systematic review of different aspects of *C. auris*; molecular epidemiology, virulence and pathogenicity, resistance, crude mortality rates and infection prevention and control in 2018 (8), but did not collate data or make interpretations regarding our outcomes of interest. Vaseghi et al. carried out a systematic overview of the literature of COVID-19 associated *C. auris* infections (20). The relevant results of the review were underlying medical conditions and medical device interventions as risk factors/predictors for *C. auris*, reflecting a highly selected group of patients.

The most frequent predisposing factors were found to be hypertension (15/996; 1.5%), followed by diabetes mellitus (12/996; 1.2%) and cardiovascular diseases (7/996; 0.7%). Central venous catheter (CVC) was found to be the most applied medical device (prevalence rate: 96%). The estimated odds ratios indicated that COVID-19 patients who received a CVC had 2.6 times the odds of catching *C. auris* co-infection. Vinayagamoorthy et al. conducted a systematic review examining the prevalence, risk factors, treatment, and outcome of *C. auris* infections in COVID-19 patients (21). They found that hypertension (17/35; 48.6%) and diabetes mellitus (12/35; 34.3%) were the most prevalent underlying conditions among COVID-19- patients with *C. auris* candidemia. Regarding risk factors, the review identified broad-spectrum antibiotic usage (35/35; 100%), ICU stay (33/35; 94.3%), mechanical ventilation (24/35; 86.6%) and CVC (28/35;80%) as the most frequently found. However, the authors found no significant differences in underlying disease and risk factors among *C. auris* infections excluding candidaemia or colonisation, and *C. auris* candidemia cases.

### Primary studies

#### Duration of C. auris colonisation

We included eight studies reporting on duration of *C. auris* colonisation (Table 4). The studies were from USA (n=5), UK (n=1), India (n=1), and South Korea (n=1), and were published between 2016 and 2023. The combined study population consisted of adult patients with a median age ranging from 51-72 years. Included participants were hospitalised in different types of hospitals and often with several underlying medical conditions. The assessment on colonisation duration was predominantly conducted within the healthcare settings, with only one study tracking patients post-discharge (22). The number of positive *C. auris* patients followed up with one or several screening tests varied from three to 75 patients among the studies. In some studies patients were lost to follow-up so the final number of patients in the included studies were three to 60 patients. Two of the studies followed less than ten patients (23;24). Follow-up time and number of screening tests taken of each patient varied between studies and among patients. Most studies followed some of the patients for at least 6 months, although follow-up time varied from a few days to 20 months.

**Table 4.** Overview of characteristics and findings from studies on the duration of C. auris-colonisation.

| Author (Year) | Country | Study period | Setting | Follow up cohort (N) | Loss to follow up (N) | Age of cohort Median years (IQR) | Follow up time<sup>a</sup> | Screening site | Screening interval | Diagnostic method | Clearance defined | Duration of colonisation |
| --- | --- | --- | --- | --- | --- | --- | --- | --- | --- | --- | --- | --- |
| Adams (2018) | USA | 2013 - 2017 | Hospitals | 38 | 2 | 72 (21 - 96) | 0 - >250 days | A, G, N, R, U, W | No standardised interval | Culture (selective agar, broth), MALDI-TOF | 2 negative cultures taken >1 week apart | Days - >200 days |
| Arenas (2023) | USA | 2021 - 2022 | Hospital | 50 | - | 68 (61 - 78) | 2 - 33 weeks | A, G | Twice weekly | PCR (rt-PCR) | Not defined | Few days - 33 weeks |
| Bergeron (2021) | USA | 2017 - 2019 | Community | 75 | 30 | 63 (51 - 70.5) | 0 - 20+ months | A, G | Every three months<sup>b</sup> | PCR (rt-PCR), Culture, MALDI-TOF | 2 negative PCR/fungal cultures taken >1 week apart | Days - 12 months<sup>c</sup> |
| Eyre (2018) | UK | 2015 - 2017 | Hospitals | 70 | 10 | 52 (42 - 64) | 6 months | A, G, N, U, W, other | Weekly and on discharge | Culture (broth, selective agar), MALDI-TOF, WGS | 2 - 3 negative screening | 2 - 3 months<sup>d</sup> |
| Jung (2020) | South-Korea | 2016 - 2018 | Hospitals | 24 | 17 | 56 (43 - 65) | 1 - 16 months | E | No standardised interval | Culture (selective agar), MALDI-TOF | Not defined | 1 - 11 months<sup>e</sup> |
| Pacilli (2020) | USA | 2016 - 2018 | Acute/long-term facilities | 51 | - | 63 (18 - 94) | 1 - 10 months | A, I | Median 35 days (28 - 84 days) | PCR (qPCR, subset), Culture (selective agar, broth) | Not defined | Up to 10 months |
| Vallabhaneni (2016) | USA | 2013 - 2016 | Hospitals | 3 | - | Not defined | Not defined | A, G, N, R | No standardised interval | Initial methods varied, WGS | Not defined | 1 - 3 months |
| Yadav (2021) | India | 2019 - 2020 | Hospital | 12 | - | 51 | 10 - 150 days | A, G, N, E | Every week | Culture (selective agar), MALDI-TOF, ITS sequencing | Not defined | 10 - 60 days |

A, axilla; G, groin; N, nasal; R, rectum; U, urine; W, wound, I, inguinal; Other, tracheostomy; E, ear; IQR, Interquartile range; PCR, polymerase chain reaction; MALDI-TOF, matrix-assisted laser desorption/ionization time-of-flight; ITS, internal transcribed spacer; WGS, whole-genome sequencing
<sup>a</sup>From first positive test; <sup>b</sup>Varied by patients; <sup>c</sup>Median time for patients to be serial negative was 8.6 months (IQR: 5.7 - 10.8). Approx. two thirds (62%) of patients colonised with *C. auris* and discharged to a community setting no longer have detectable *C. auris* colonisation; <sup>d</sup>Median duration of colonisation among patients was 61 days when two consecutive negative screening results were used to define clearance of colonisation and 82 days when three consecutive negative results were used; <sup>e</sup>Seven patients had follow-up culture results. Two patients had positive culture at 8 months after first isolation and one patient at 11 months.

Three studies lacked a pre-defined, standardised interval for the follow-up screening (23–25). Although the other studies had defined screening intervals, the time frames varied among the patients involved. Colonisation clearance was not clearly defined in five of the studies (23;24;26-28). All the studies, except one, obtained follow-up samples from multiple sites, primarily the groin and axilla. The exception conducted screenings solely from external ear discharge (24).

The absence of predefined, standardised protocols for research into colonisation duration, including set time intervals and the number of tests to conduct, coupled with the lack of unified definitions for spontaneous decolonisation, complicates the interpretation of duration of *C. auris* colonisation.

Seven studies looked at the length of colonisation among patients in hospitals (16;23-28). The duration of colonisation varied between studies and patients, ranging from days to years, with long-term colonisation of a few months to over a year being commonly observed. Eyre et al. followed 60 patients for up to 6 months with weekly screening and at discharge (16). They found the median duration of colonisation to be 2-3 months. Clearance was defined as two or three consecutive negative cultures. Pacilli et al. followed 51 patients with screening every week and found the duration of colonisation to be up to 10 months in some patients, but were unable to ascertain their definition of colonisation clearance (27). Adams et al. analysed follow-up samples from 38 patients, collected at least one week apart, and observed a duration of colonisation ranging from days and up to >200 days (25). Bergeron et al. assessed colonisation among patients post discharge with a follow-up time ranging from 0 to 20 months, taking samples every three months. They found that 62% of the patients did not remain colonised in the follow-up cultures after discharge to the community. They defined ‘serial negative’ as two consecutive negative PCR or fungal culture results. The median time for patients to become ‘serial negative’ was 8.6 months (interquartile range: 5.7-10.8) (22). However, across studies, persistent colonisation for up to a year post-discharge was still common, suggesting that clearance is inconsistent and may be influenced by screening frequency or methodology.

We extracted data on the diagnostic methods used in each study, documenting the sequence in which they were applied. These methods included culture-based approaches (selective agar, broth) with or without Matrix-assisted laser desorption/ionization time-of-flight (MALDI-TOF) confirmation and molecular methods such as polymerase chain reaction (PCR). Three studies incorporated PCR at some stage in the diagnostic process (22;26;27), but given the substantial heterogeneity in reported colonisation durations, no apparent differences were observed compared to studies that relied solely on culture-based methods. Given the known challenges in culturing *C. auris*, studies incorporating molecular identification at the initial stage may have had a higher detection sensitivity than those relying exclusively on culture. However, no study directly compared the two approaches in a way that allowed for conclusions about differences in measured colonisation duration, and as this review focused on colonisation duration rather than diagnostic accuracy, studies solely concerning microbiological methods were not included.

A critical question regarding recommendations for screening, as well as other infection control measures, is whether individuals previously diagnosed with *C. auris* should be considered permanent carriers or if negative test results can lead to the cessation of infection control measures. Multiple studies have demonstrated that patients can experience one or more negative screening result, prior to a subsequent positive result (26–28). Pacilli et al. observed that 55% of the patients who had at least one negative screening result later tested positive in a subsequent sample (27). In two separate studies, Arenas et al. and Bergeron et al. found that 34% and 25% of colonised patients, respectively, received a positive result after at least one negative result, with the time interval between screenings varying between patients in both studies (22;26). Arenas et al. used rt-PCR, while Bergeron et al. combined fungal culture and rt-PCR, which may have contributed to the differences in reported rates of intermittent negative results (22;26). Hence, negative test results for a colonised patient should be interpreted with caution when considering discontinuation of appropriate infection prevention and control precautions in healthcare settings. Such a careful approach is supported by international guidelines. The CDC does not recommend routine reassessments for *C. auris* colonisation, as intermittent negative results have been observed and patients may remain colonised long-term despite negative screenings (29). An expert meeting organised by the International Society for Antimicrobial Chemotherapy in 2019 recommended maintenance of infection control measures until discharge and to flag the patient chart for at least 1 year after the first negative screening culture (30).

### Risk of secondary cases

We included 15 studies that reported on the risk of secondary cases of *C. auris* (Table 5). These were conducted in Australia, Canada, Colombia, India, Italy, Saudi Arabia, Republic of Korea, Spain, UK and USA. Studies were published between 2017 and 2024. The studies reported on outbreak investigations and surveillance screening. The study settings were hospitals, mainly ICUs, with two studies conducted in nursing facilities with ventilator-capability (31;32). Index *C. auris* cases, here defined as the primary cases that were followed for potential onward transmission, ranged from one to 50 adults, depending on study design, number of hospitals included, and the study period. The total number of tested (investigated) persons ranged from 17 to 960. Tested persons were mainly patients, with seven studies including healthcare workers (HCWs) and/or family members and visitors.

**Table 5.**
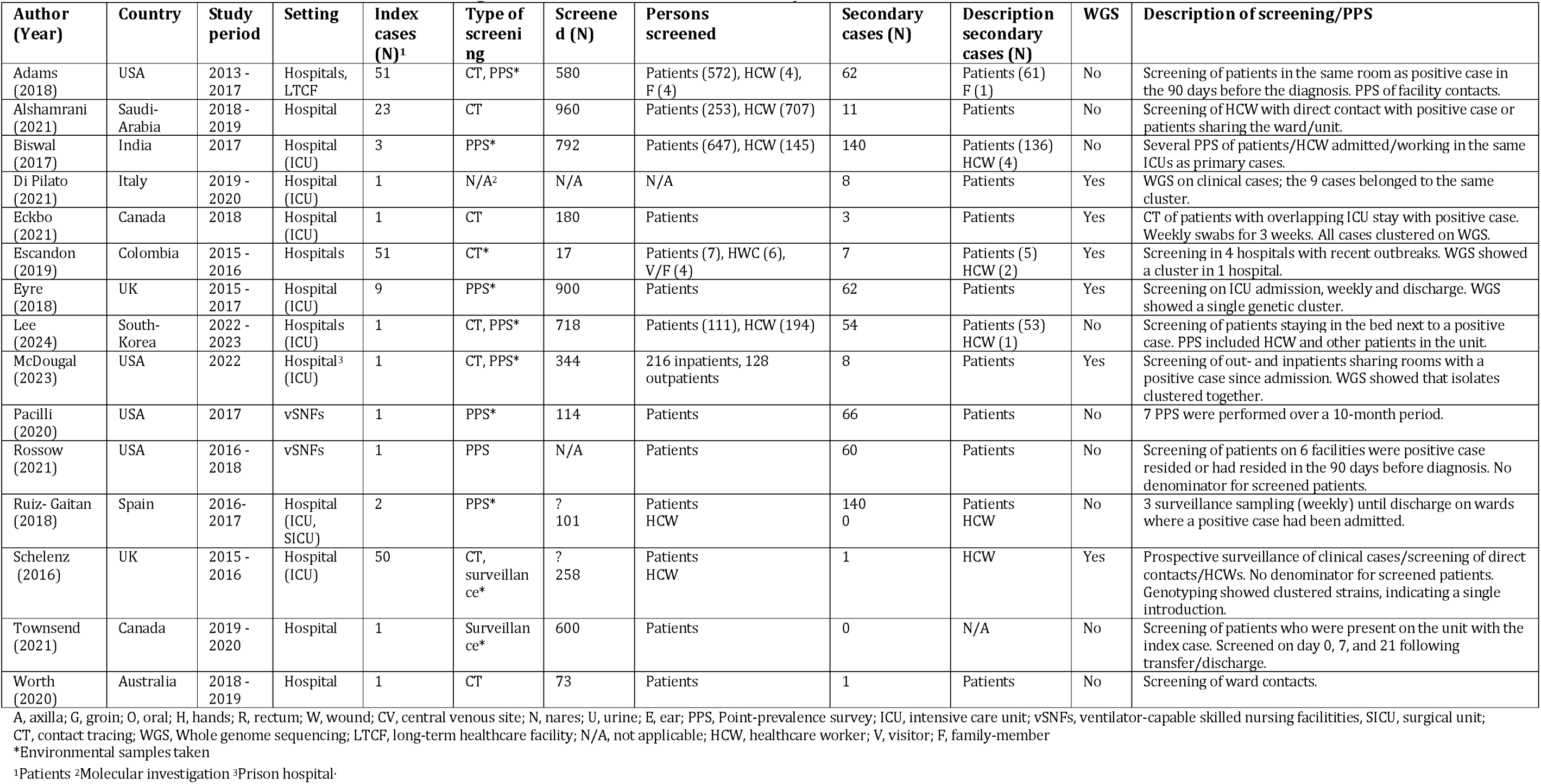
Overview of characteristics and findings of studies on risk of secondary cases of C. auris.

While seven studies reported more than 50 secondary cases, three studies observed zero to three secondary cases, despite testing between 180 and 600 patients (33). Secondary cases were mainly reported in patients. Due to differences in test strategies, follow-up periods, and number of defined primary cases, we could not calculate a secondary attack rate. Some studies did not report the total number of tested individuals. However, as the proportion of positive cases out of tested patients were >40% in some studies, a high risk of secondary cases has been documented in outbreak settings (27;34). Several studies showed that a high number of patients were already colonised shortly after the outbreak was recognised, but in some situations transmission was limited to a small number of patients, and the proportion of positive cases out of those tested were <2% (33;35-37). The reason for this may be multifactorial – i.e., transmission depend on the setting, the population, when the outbreak was detected, the amount of environmental contamination, and the infection control measures implemented.

Four studies detected positive cases among HCWs, identifying between one and four positive individuals from a pool of six to 258 tested persons (34;38-40). Schelenz et al. found one positive HCW out of 258 tested in an outbreak where HCWs had been caring for a patient colonised with *C. auris* (40). Escandon et al. found two HCWs that had positive hand/groin samples out of six tested (34). The two last studies found one and four HCWs where samples from their hands were positive, respectively, when performing point-prevalence surveys (PPS) in the units where there had been a positive case (38;39). One out of two studies testing family members found one positive family-member (25). The selected studies discovered minimal positive instances among HCWs, primarily featuring yeast present on their hands, even though extensive testing was conducted during outbreaks. Several authors have postulated that HCWs could potentially contribute to transmission chains due to transient hand contamination. However, to affirm this as a prevalent route of transmission, additional research is required.

Six studies reported results from PPS or surveillance screening (16;27;31;33;38;41). The authors reported testing of 101 to 900 persons and found 0 to 140 positives. Four studies did a combination of PPS, surveillance, and contact tracing (25;39;40;42). They tested 258 to 718 persons and found 1 to 62 positive cases. We were not able to find a denominator in three of the studies, and several studies had defined up to 50 primary cases in different hospitals and did surveillance screening on the wards where the cases were admitted (25;34;35). Six out of 15 studies did WGS, confirming genetically distinct clusters.

Not all studies differentiated between direct and indirect exposure as sources of secondary cases. Some studies clearly defined direct exposure and contact tracing, while in several others, the results were presented alongside the number of detections of *C. auris* in broad surveillance surveys. In a comprehensive approach, four studies carried out contact tracing, testing patients and HCWs with known exposure to a confirmed case. This exposure included direct contact, overlapping stays in the ICU, or other connections within the same ward (34–37). These studies tested 17 to 960 exposed persons and found one to eleven positive cases. Lee et al. concluded that the proportion of secondary cases of *C. auris* from close contacts was relatively high (25%) (39). However, Rossow et al. did not find an association between *C. auris* colonisation and residing in a room with a positive case (31).

To determine the transmissibility of *C. auris*, several factors are crucial, including the microbe’s inherent virulence, host susceptibility, the type and duration of contact, and persistence in the environment. Schelenz et al. conducted a root cause analysis and discovered that the minimum duration of contact with a positive case or a contaminated environment for acquiring *C. auris* was ≥4 hours. However, they were unable to identify a single specific source of transmission (40). Existing expert recommendations have emphasised the importance of prompt screening and infection prevention measures in healthcare settings to prevent nosocomial transmission of *C. auris* (43), as is supported by international guidelines. ECDC recommends prompt and robust measures if *C. auris* is detected in healthcare facilities including testing of close contacts (44), while CDC recommends testing patients sharing the same room, unit, or other care areas with a patient with *C. auris* (45). While these guidelines or their adherence have proven insufficient to prevent both major outbreaks and increasing prevalences (46), experiences from both the United Kingdom and Germany demonstrate that successful containment of such outbreaks is possible (16;47).

The included studies used different definitions of screening, and several studies presented results from contact tracing alongside the number of detections of *C. auris* in broad surveillance surveys or screening. Caution should be exercised when directly incorporating these results into decisions regarding admission screening recommendations. Conversely, numerous studies have reported secondary cases from screening patients who either shared a room as a positive case or occupied the neighbouring bed. This could support that an individual with a high degree of exposure is likely to have a high risk of becoming infected and may warrant their inclusion in screening protocols. Moreover, occupying the same environment as a *C. auris* case can potentially lead to colonisation or infection.

### Associated factors of **C. auris** infection or colonisation

A total of 112 studies reported on factors associated with colonisation or infection with *C. auris* (Table 3). These studies were conducted in 30 different countries from all inhabited continents, with two studies including more than one country and two not specifying the country of sampling. Most studies primarily focused on detailing patients’ medical profiles or conditions without a control group and described critically ill patient populations or those with severe underlying medical conditions. ICUs were the predominant setting for these studies. Fourteen of the included articles included a control group and investigated factors associated with either colonisation or infection *C. auris* (Table 6) (31;48-60). These studies were predominantly small-scale (n<100) and based on outbreak investigations in heterogeneous populations. Most used bivariable screening to identify potential explanatory covariates, which were then included in multivariable regression models using stepwise selection techniques. However, this methodological approach may limit the validity of the identified factors due to several well-documented issues such as the multiple comparisons problem, exacerbated by the small sample sizes, and a lack of theoretical reasoning underpinning the modelling choices (61;62). Therefore, when reviewing these studies, counting how many times a factor has been found statistically significant in the included studies would not lead to robust inferences about their relative importance.

**Table 6.** Overview of characteristics and findings of studies on associated factors (control group) for C. auris infection or colonisation.

| First author (year) | Country | Study period | Outcome type | Exposure group | Control group | Exposed (n) | Control (n) | Risk factors (authors' conclusions) |
| --- | --- | --- | --- | --- | --- | --- | --- | --- |
| Adam (2019) | Kenya | 2010-2016 | Candidaemia | *C. auris* candidaemia | Other candidaemia | 77 | 124 | Carbapenem use, critical care unit, presence of CVC |
| Allaw (2022) | Lebanon | 2020-2021 | Infection (bloodstream, deep tracheal aspiration, urine, wound) | *C. auris* positive with or without severe covid-19 | *C. auris* negative with or without severe covid-19 | 56 | 130 | qSOFA, length of stay in hospital |
| Alvarez-Moreno (2023) | Colombia | 2016-2017 | Mortality (crude 30 and 90 day) | *C. auris* candidaemia | Other candidaemia | 22 | 52 | Previous use of fluconazole, HIV |
| Caceres (2020) | Colombia | 2015-2016 | Candidaemia | *C. auris* candidaemia | Other candidaemia | 40 | 50 | BSI: $\geq 15$ days of pre-infection ICU stay, evidence of severe sepsis, and diabetes mellitus |
| Farooqi (2020) | Pakistan | 2015-2016 | Colonisation or infection, confirmed or suspected | *C. auris* confirmed or suspected | Other admitted patients matched on age and sex | 30 | 62 | History of surgery 90 days before diagnosis, admission through the ED and having chronic kidney disease |
| Moin (2021) | Pakistan | 2020 | Candidaemia | *C. auris* | Other candidaemia | 4 | 22 | Prior antifungal exposure |
| Parak (2022) | South Africa | 2015-2018 | Candida spp. Isolated | *C. auris* | *C. albicans* and *C. glabrata* | 45 | 90 | Indwelling devices and previous antibiotic exposure |
| Rossow (2021) | USA | 2016-2018 | Colonisation | *C. auris* | Controls from the same facility without *C. auris* | 60 | 218 | Specific invasive medical devices, recent antimicrobial use, recent hospitalization, and colonisation with other MDROs |
| Ruiz-Gaitan (2019) | Spain | 2016-2017 | Colonisation or candidaemia | *C. auris* | Concurrent controls without *C. auris* | 114 | 114 | The presence of CVC, parenteral nutrition and mechanical ventilation |
| Sayeed (2020) | Pakistan | 2014-2017 | Candidaemia | *C. auris* | Other candidaemia | 37 | 101 | Prior history of surgery, prior antifungal exposure, and MDR bacteria isolation |
| Shastri (2020) | India | 2016-2017 | Candidaemia | *C. auris* | Other candidaemia | 42 | 66 | Underlying respiratory or neurological diseases. Presence and use of CVC for long period, prolonged ICU stay prior to candidaemia, invasive ventilation, and broad-spectrum antibiotic usage |
| Simon (2023) | USA | 2016-2020 | Candidaemia | *C. auris* | Other candidaemia | 83 | 113 | Admission from nursing homes, prior colonisation with *C. auris*/multidrug-resistant bacterial organisms, extensive use of antimicrobials concomitant bacterial infections with gram-negative and multidrug-resistant organisms. |
| Taori (2019) | UK | 2016-2017 | Clinical infection | *C. auris* infection | *C. auris* colonisation | 8 | 6 | ICU stay, long hospital stays, presence of CVC, haemodialysis, and prior antifungal therapy |
| van Schalkwyk (2019) | South Africa | 2016-2017 | Candidaemia | *C. auris* | Other candidaemia | 794 | 5875 | Prior systemic antifungal drug therapy, presence of a CVC, admission to a private-sector facility, older age, longer hospitalization before the first positive blood culture, prior hospitalization within the past year, admission to an ICU during the current hospital stay, and HIV seropositivity. |

Exposure to the microbe is necessary for colonisation to occur. However, there have been very few studies investigating such exposure specifically, even though existing guidelines may target persons assumed to be exposed for instance through contact with healthcare system in endemic regions (63;64). We have only found one case series and one study examining the risk associated with travel and contact with healthcare abroad. In the case-series by Hamprecht et al., *C. auris* was found in seven patients in Germany between 2015 and 2017, six of whom had previously been treated in healthcare centres in the Middle East, Asia, Africa, or the United States (65). In a study of healthy travellers by Turbett et al. (66), 94 individuals were screened pre- and post-travel using self-collected axillary-inguinal groin swabs for *C. auris*. Travel destinations included all major regions of the world, with eastern Africa (31/94, 33%) being the most common region visited, and leisure being the predominant reason for travel (71/94, 76%). No *C. auris* was isolated from the samples. Few travellers (5%) reported providing or receiving medical care while traveling, indicating a low incidence of contact with healthcare systems during travel. Although the basis for inferences is sparse, the other studies align with the findings of these two studies, highlighting high-risk healthcare settings, particularly ICUs, as common sites of acquisition. However, recent studies confirm that *C. auris* may also persist in non-healthcare environments, such as stored fruit, where exposure to agricultural fungicides could contribute to antifungal resistance (67).

Prior reviews have identified correlations between *C. auris* infection with certain general factors, such as male gender along with severe underlying conditions such as immunosuppression, diabetes, and chronic kidney disease (8;68). Additionally, the presence of invasive devices and procedures is consistently cited as a risk factor (8;68;69). These general factors may be explained by our finding that most studies were conducted in critical care unit settings, as these patients are often implicated in hospital outbreaks or contract severe (and resistant) healthcare-associated infections, regardless of the specific causative agent. Some factors, like prior antifungal exposure, might be more specific, but it remains uncertain whether this selects for *C. auris* specifically or de-selects for other yeasts (8;68). Broad-spectrum antibiotics may also play a role by eliminating competing microbiota, thereby facilitating *C. auris* colonisation (68;69).

Colonisation with other MDROs could indicate shared risk factors between these multidrug-resistant pathogens (68). Past hospitalisation may also capture the statistical association of this shared risk set, and its inclusion in models should be carefully evaluated to first determine whether it acts as a mediator or confounder in the exposure-outcome relationship. The inclusion of these and other covariates without such a consideration may result in an attenuation of effect sizes, or even lead to incorrect conclusions.

The majority of studies which included a control group compared outcomes between patients with *C. auris-* candidaemia and those with other types of candidaemia. The research by van Schalkwyk et al. stands out due to its large scale, encompassing a national survey conducted over two years in an endemic region (60). This study evaluated a predefined hypothesis, among others, demonstrating a substantial increase in *C. auris* candidaemia. Rossow et al. concentrated their research on individuals colonised by *C. auris*, comparing them to non-colonised controls from identical exposed cohorts. They identified factors associated with colonisation, including the use of specific invasive medical equipment, recent use of antimicrobials, recent hospitalisations, and colonisation with other MDROs (31). When evaluating factors associated with infection or colonisation that are relevant to screening, the crucial variables are those helping us differentiate between incoming hospital patients who are at risk of being colonised and those with minimal risk. While comparisons with other candidaemias can highlight distinct features of *C. auris*, they offer limited insights into screening strategies for colonisation.

### Colonisation sites

Twenty-nine studies reported on colonisation sites for *C. auris* and their positivity rates based on predetermined sampling materials (Table 7). The studies were conducted in 11 different countries, most of them in the USA (N=12), in the period 2013 to 2023. Armstrong et al. and Escandon et al. reported on the same materials sampled from patients, but not from HCWs (34;70). The studies varied in designs and populations, and revealed diverse patterns, prevalence, and positive sites for the detection of *C. auris*. Differences in diagnostic methods could be a source of the variation observed across studies. Among the 29 studies included, the majority (N=23) relied exclusively on culture-based methods, using either selective agar or broth enrichment, often followed by MALDI-TOF confirmation. Five studies incorporated PCR at some stage in the diagnostic process (63;71-74), and one study exclusively used PCR (75). While we did not observe any systematic differences in reported positivity rates between studies using PCR and those relying solely on culture-based methods, variations in diagnostic approaches may contribute to the heterogeneity seen in colonisation site detection.

**Table 7.** Overview of characteristics and findings of studies on colonisation sites for C. auris.

| First author (Year) | Study period | Country | Setting | Population | Type of screening | Number of screened persons (or samples) | Screening sites | Diagnostic method | Total positive N (%) | Positive body site |
| --- | --- | --- | --- | --- | --- | --- | --- | --- | --- | --- |
| Adams (2018) | 2017 - 2017 | USA | Hospital, LTCF, vSNF | Patients, HCW and family members | CT and PPS | 346 patients | A, G, N | Culture (selective agar, broth), MALDI-TOF | 36 | A-G 13/36 (36%), N 9/36 (25%), A-G-N 14/36 (39%) |
| Alshahrani (2023) | 2020 – 2022 | Saudi Arabia | Hospital | Patients | On admission, CT | N/A | A, G, N (W, IDS, U, etc) | Culture (selective agar), MALDI-TOF | 46 | A 6 (11.3%), U 16 (30.2%) * |
| Armstrong (2019) | 2015-2016 | Colombia | Acute care hospitals, | Adult and pediatric patients | Different body sites of positive *C. auris* patients | 7 patients | A, G, bilateral N, E, O, R | Culture (broth, selective agar), MALDI-TOF | 5 | P1 (BSI): 1/11 R, P2 (BSI): 0/10, P3 (BSI): 7/11 E, A, N, R, fecal material, P4 (UVI): 1/10 G, P5 (sputum): 1/11 G |
| Biswal (2017) | 2017 | India | Hospital | Patients, HCW | On admission, HCW | 647 patient samples<br>145 HCW samples | A, G, O, R + H (HCW) | Culture (selective agar), MALDI-TOF | 136 (4) | A 72/196 (36.7%), G 36/206 (17.4%), R 18/118 (15.2%), O 10/95 (10.5%), HCW hands 4/145 (2.8%) |
| Briano (2022) | 2020-2021 | Italy | Hospital | ICU patients | On admission, weekly and on discharge | N/A | Combined A-G | MALDI-TOF (no culture details) | 157 | S 146/157 (93%), U 38/157 (24%), RT 77/157 (49%), multisite 48/157 (50%) |
| Chow (2018) | 2013-2017 | USA | Acute care facility | Patients | CT, PPS | N/A | A, G (some N, W) | Culture (broth), MALDI-TOF | 60 | Mixed infection and colonisations |
| Corcione (2022) | 2021-2022 | Italy | Hospital | ICU patients | CT, PPS | N/A | A, G, N, T | Culture (selective agar), MALDI-TOF | 8 | S 5/8 (%), U 2/8 (%), RT 1/8 (%) |
| De Melo (2023) | 2021-2022 | Brazil | Hospital | Patients | CT | N/A | A, G | Culture (selective agar), MALDI-TOF, PCR | 7 | 7/7 A-G |
| Escandon (2019) | 2015-2016 | Colombia | 1 acute care hospital. 2 adult hospital, 1 pediatric hospital | Adult and pediatric patients | Different body sites of positive or suspected *C. auris* patients and HCW | 17 | A, G, bilateral N, E, O, R + HCW hands | Culture (broth), MALDI-TOF, WGS | 5 patients<br>2 HCW | H 2/2 HCW G 1/2 HCW |
| Eyre (2018) | 2015-2017 | UK | Hospital | Neuroscience ICU patients | On admission, weekly and on discharge | 900 patients | A, G, N, U (W, T) | Culture (broth, selective agar), MALDI-TOF, WGS | 60 | A 22/60 (37%), G-U 21/60 (35%), multisite 17/60 (28%) |
| Goulart (2023) | 2021 – 2023 | USA | Trauma center | Patients | PPS | 91 patients | A, G | PCR, Culture | 5 | Not specified which site |
| Lee (2024) | 2022 | South Korea | Hospital | ICU/CCU Patients and HCW | CT, PPS, HCW | 111 patients<br>194 HCW | Bilateral A, bilateral G | Culture (selective agar), MALDI- | 53 patients and 1 HCW | Results mixed for patients with infection and colonisation. HCW positive hands/pockets |
|  |  |  |  |  |  |  |  | TOF |  |  |
| Leonhard (2024) | 2023 | Netherlands | University medical center | Patients | On admission | 199 patients | A, G (IDS, U etc) | PCR, Culture (selective agar, broth), MALDI-TOF | 1 | A-G 1/1 (100%) |
| McDougal (2023) | Not specified | USA | Prison hospital | Patients in prison health care | CT, PPS | 344 patients | Combined bilateral A-G | Culture (selective agar), MALDI-TOF | 8 | A-G 8/8 (100%) |
| McPherson (2020) | 2019 | USA | Hospital (long term transitional care) | Pediatric patients | PPS | 25 patients | Combined bilateral A-G | PCR (rt-PCR), Culture (not specified) | 0 |  |
| Mulet Bayona (2020) | 2017 | Spain | Hospital | ICU patients | On admission, follow-up, PPS | N/A | N/A | Culture (blood culture, selective agar), MALDI-TOF | 35 | 9 A-R-P, 24 A-R and 2 P |
| Piatti (2022) | 2021 | Italy | Hospital | ICU patients | On admission, on discharge | 384 patients | A, G, E, IS, R (some) | Culture (selective agar), MALDI-TOF |  | S 105/384 (27,3%), R 50/86 58,1%), 69/77 (89,6%) |
| Proctor (2021) | 2019 | USA | Skilled nursing facility |  | PPS | N/A | A, G, N, E, PS, TW, PF, B, TO, T | Culture (selective agar, broth), MALDI-TOF | 49 | N 42,9%, foot 35,7%, PF 40,4% |
| Rowlands (2023) | 2017-2019 | USA | 2 ventilator units nursing homes and 1 hospital (ventilator/pulmonary unit, ICU and cardiac care unit) | Patients | On admission | 2062 patients | Combined bilateral A-G, bilateral N | PCR (rt-PCR) | 188 | A-G 93/188 (49.5%), N 32/188 (17.0%), A-G-N 61/188 (32.4%) |
| Ruiz-Gaitan (2018) | 2016 – 2017 | Spain | Tertiary care hospital | Patients and HCW | PPS | N/A | S, TH, R, H (HCW), E | Culture (selective agar), MALDI-TOF, ITS sequencing | 140 | Results mixed for patients with infection and colonisation |
| Schelenz (2016) | 2015 – 2016 | UK | Hospital Trust | ICU patients and HCW | CT | N/A | A, G, N, R, W, U, IDS | Culture (selective agar), MALDI-TOF | 50 | Results mixed for patients with infection and colonisation. N (HCW) 1/258 |
| Sharp (2021) | 2017 – 2018 | UK | 8 adult ICUs in different hospitals | ICU patients | On admission | 921 | A, G, N, TH, PS, R, CU | Culture (selective agar), MALDI-TOF | 0 |  |
| Southwick (2022) | 2016 - 2017 | USA | Different Health care facilities | Patients | CT and PPS | 1668 patients | Combined A-G, N | Culture (agar), PCR | 114 | A-G 92/114 (81%), N 73/114 (64%), A-G-N 68/114 (60%) |
| Townsend (2021) | 2019 – 2020 | Canada | Acute care community hospital | Patients | On admission | N/A | Bilateral A, G, N | Culture | 1 |  |
| Tsay (2017) | 2016 - 2017 | USA | Healthcare facilities | Patients | CT | 390 patients | Combined A-G, N (some) | Cannot be determined, WGS used on isolates | 45 | A-G 45/45 (100%) |
| Turbett (2022) | 2019 - 2020 | USA | Travel to Africa, Asia, USA, Europe | Travellers | Before and after travel | 94 travellers | Bilateral A, bilateral G | Culture (broth, selective agar), MALDI-TOF | 0 |  |
| Vallabhane ni (2017) | 2013 - 2016 | USA | Hospitals | HCW | HCW hands | 52 samples | Hands | Initial methods varied, WGS | 0 |  |
| Yadav (2021) | 2019 - 2020 | India | Chest hospital | Patients | On admission<sup>b</sup> | 32 patients | A, G, N, E | Culture (selective agar), MALDI-TOF, ITS sequencing | 12 | G 9/12 (75%), N 5/12 (45%), E 4/12 (33%) |
| Zhu (2020) | 2016 - 2018 | USA | Hospitals, nursing homes, hospice, LTACH | Patients | 1) Surveillance and<br>2) different body sites of positive *C. auris* patients | 11035 samples | A, G, N (R, other) | Culture (pre-cultured isolates), MALDI-TOF | 1) 931 samples (350 patients)<br>2) 298 | A-G bilateral 178/222 (80%), N bilateral 125/215 (58%), A-G-N bilateral 106/103 (100%), A unilateral 10/20 (50%), G unilateral 10/20 (50%), N unilateral 6/14 (43%), W 4/11 36%, R 4/7 (57%), E 4/6 (67%), S 1/1 (100%) |

PPS, Point- prevalence survey; CT, contact tracing; LTCF, long-term healthcare facility; vSNF: ventilator Skilled nursing facility; N/A, not applicable; HCW, healthcare worker; A, axilla; B, buccal mucosa; CU, catheter urine; CV, central venous site; E, ear; G, groin; H, hands; IDS: Indwelling device site; IS: Inframammary sulcus; N, nares; O, oral; P, pharyngeal; PF: palm and fingertips; PS: perianal skin; R, rectum; RT, respiratory tract; TH; throat; TO: tongue; TW: toe web; U: Urine; W, wound; PCR: polymerase chain reaction; MALDI-TOF: matrix-assisted laser desorption/ionization time-of-flight; ITS: internal transcribed spacer; WGS: whole-genome sequencing
\* Additionally, patients were positive in the thigh (n=5), anus (n=5), arm (n=4), penis (n=3), hip (n=3), buttock (n=2), leg (n=2), neck (n=1), tissue (n=1) wound (n=1) nail (n=1), foot (n=1) area.

The axillae and groin regions were the most common sites of *C. auris* colonisation, but also the body sites from which samples were most frequently taken. Positivity rates for screening sites in *C. auris* positive patients ranged from 36% to 100% for combined sampling of axilla and groin, 11.3% to 50% for axillae alone and 17.4% to 75% for groin separately (28;38;76). Colonisation of the nares was also observed, with positivity rates ranging from 17% to 64% (25;28;74-77). Piatti et al. used swab specimens from the skin of axilla, groin, auricular area and inframammary sulcus as screening sites and found a 27.3% (105/384) prevalence of skin colonisation of admission screenings in Italy (78). Additionally, positive cases were detected in other body sites such as rectum, oral cavity, respiratory tract, ear, urine, and wounds (76;79). Three studies reported separate results for the colonisation of *C. auris* on the hands of HCWs (34;38;39). Biswal et al. found that out of 145 screened HCWs, four (2.8%) tested positive. Whether the samples were reported as being taken bilaterally, in a composite swab, or individually varied between studies. ECDC recommends the axilla and groin as screening sites, as well as other sites (urine, wounds, catheter exit sites, throat etc.) if clinically relevant or indicated (44) and US CDC recommends bilateral screening of axillae and groin (80). Pan American Health Organization (PAHO) recommends screening the axilla, oropharynx, nostrils, groin, urine and rectum, but if it is not feasible to collect samples from all sites, they recommend at least pooled samples from the groin and axilla (81).

A literature review by the United Kingdom Health Security Agency draws attention to the study by Adams et al. where the addition of a sample from the nostril increases the probability of detection (25). However, Rowlands et al. and Southwick et al. reported lower positivity rates when axillae, groin and nares were combined as screening site than axillae and groin alone (74;75). Studies employing screening methods where the pre-test probability of a positive screening test is higher (contact tracing, PPS) also consistently identified the axillae and groin as primary sites for *C. auris* colonisation. Southwick et al. found that among 1668 patients, 144 (7%) were positive for *C. auris* colonisation, and positive results were predominantly observed in composite samples from the axilla and groin (81%), followed by the nares (64%) (74). Zhu et al. conducted an outbreak investigation in 2020 and included 11,035 samples of which 931 (8.4%) were positive for *C. auris* (76). The findings revealed that 80% (178/222) of positive samples were positive for *C. auris* from the axilla-groin (tested with a composite swab) and 58% (125/215) in the nares. They also found that if the nares are colonised, they harbour a relatively higher amount of *C. auris* than does the axilla or groin. Notably, when considering the combination of axillae-groin-nares, the positivity rate approached 100%. In other words, sampling from the axilla and groin bilaterally of patients at risk, as well as the nares, may find justification. While the cost-effectiveness of sampling from the nares warrants evaluation, the potential for increased detection rates may justify its inclusion, given the significant concerns surrounding over the spread of *C. auris*.

A limited number of the included studies aimed to discern the body sites where *C. auris* resides by testing various areas on infected patients. In only three studies, multiple body sites in patients with confirmed *C. auris* infection were screened (70;76;78). Armstrong et al. included five positive *C. auris* patients and screened each patient 10-11 times at different sites (70). There was significant variability in the body sites where *C. auris* was detected in screening. Zhu et al. reported on 298 microbiological samples where individuals had an infection, but the results reporting positive body sites are presented in combination with results from a surveillance survey, precluding data extraction regarding positive patients alone (76). Piatti et al. conducted a study using skin and rectal swabs to screen for the presence of *C. auris* on all patients admitted to at-risk units (78). Upon analysing patients with confirmed *C. auris* infection, the prevalence of colonisation in both skin and rectal areas was found to be nearly identical. Additional evidence suggests that screening multiple body sites improves detection rates (82).

A few of the included studies also tested wounds, tracheal secretions, and catheter urine and detected *C. auris.* Invasive devices have been found to be a factor associated with colonisation, although this may only apply to critically ill patients. Furthermore, some studies contain description of screening indwelling devices when present, but whether it was tested from these locations is uncertain, and no results are reported from this sample material (40;63;79).

However, there are different definitions in the literature regarding whether urine as a specimen should be considered colonisation or infection depending on clinical presentations/symptoms. The studies incorporated, which include description of urine samples, do not provide detailed descriptions. The same may apply to wounds, although Zhu et al. consider wounds as a source of colonisation (76).

### Strengths and limitations of this review

Our review has several strengths. First, we employed a systematic and reproducible search with a comprehensive strategy, and a thorough review of included studies. We chose to do a broad search for all articles on *C. auris* and have likely been able to identify all literature relevant to our outcomes of interest. Furthermore, findings were presented in the context of all existing studies, providing the most up to date overview yet. Finally, our systematic approach avoids some of the pitfalls and biases of purely narrative reviews without systematic searches. While a narrowing of the inclusion criteria could have addressed some of the heterogeneity issues we revealed, such an approach would perhaps exclude a significant proportion of the current literature. We opted for a narrative summary of our findings to provide the most comprehensive overview of this broad research field, as the heterogeneous nature of the included studies precluded a meta-analysis or other quantitative synthesis. A formal quality assessment was not performed, though this limitation is somewhat mitigated by the high variability in study designs and settings. The open nature of our research question, however, introduces a risk of creating a less focused review, which may not allow for a full assessment of the strength of specific evidence.

Additionally, without a quantitative synthesis, the findings lack the weighted perspective that a meta-analysis could offer in highlighting patterns across studies.

## Conclusion

Our systematic review summarises several key findings regarding the associated factors of colonisation/infection, the duration of colonisation, the potential for secondary cases, and possible colonisation sites of *C. auris*. However, our primary finding was that in screening for *C. auris*, several major knowledge gaps remain. Most studies reviewed described circumstances or outbreaks that arose unexpectedly, and lacked pre-developed protocols and a clear study aim that could be effectively addressed with the selected methods. The publication bias introduced by such a circumstantial body of literature must be acknowledged. If targeted screening of high-risk populations is chosen as a strategy to prevent establishment of *C. auris*, it remains unclear who should be screened, and there is no robust evidence either for or against current practices. Identifying individuals likely to have been exposed to *C. auris* remains the most critical factor in such guidelines, as highlighted also by others (68). Screening should continue to be performed in the axillae and groin, but considering additional sites like the nares could increase sensitivity, although there is conflicting evidence. Furthermore, routine screening of HCWs appears unnecessary unless there is clear evidence of direct exposure to contaminated settings. Given the lack of evidence that spontaneous decolonisation is a common occurrence among patients, it might be prudent to view exposed patients as potentially persistently colonised as a precaution.

Moving forward, the knowledge gaps we have identified herein should be addressed. There is an urgent need for primary research that methodically outlines the efficiency of current screening programs, including their sensitivity and specificity. This research should include findings from different anatomical sites and comparisons with composite swabs. Notably, given the substantial heterogeneity in the reported outcomes, it was difficult to identify any systematic difference between culture-based and molecular methods in the included studies. However, as challenges in detecting *C. auris* using culture alone are well-documented and recent studies have highlighted discrepancies between these methods, care should be taken in future research to ensure methodological consistency and accuracy (63). A suggested screening protocol advocates for using the most sensitive and specific method, PCR, initially, followed by culturing, an approach that appears promising (63). To determine colonisation duration, it is crucial to register positive patients and systematically monitor them at pre-set intervals over a specified period to delineate the natural timespan of *C. auris* colonisation. Furthermore, it is crucial to map how the hospital environments act as a vector in the transmission of *C. auris*. This not only includes the surfaces of the hospital, its ventilation, water and sanitation systems, but also the surrounding nature and animals. The emergence of *C. auris* fundamentally pertains to One Health, and this multidisciplinary approach should remain the predominant for addressing *C. auris* establishment in healthcare settings. Finally, the study of factors associated with *C. auris* colonisation should focus on identifying factors able to discriminate high-risk patients that should be screened in larger cohorts that are representative of all patients in healthcare settings. In addition to factors that may be coded from the medical charts, history of travel and contact with healthcare in endemic regions should be included in such studies. It is crucial to acknowledge that the most significant “risk factor” for contracting *C. auris* is exposure to the microbe itself. Such research initiatives as mentioned here could greatly bolster our comprehension, thus aiding the formulation of more effective screening guidelines and infection control measures for *C. auris*. In the absence of these research initiatives, however, the success of *C. auris* screening is more likely if it is grounded on medical theory, current knowledge of yeast microbiology, and existing literature on screening against other similar microbes.

## Data Availability

All data produced in the present work are contained in the manuscript.

## Data Availability

All data produced in the present work are contained in the manuscript.

## Acknowledgements

We extend our thanks to all colleagues at the Norwegian Institute of Public Health (NIPH) and Statens Serum Institut (SSI) who have engaged in discussions on this topic, particularly those involved in the project to develop updated recommendations to contain and prevent the spread of resistant microbes in our countries.

## Author contributions

LEØ, RR, and MM conceptualized the study and designed the methodology with JPWH. Data curation was performed by RAT. LEØ, MM, and ASD conducted the investigation and formal analysis. Project administration was overseen by MM and HMEV, while JPWH provided supervision. The original draft was written by ASD, LEØ, and MM. All authors reviewed and edited the manuscript.

## Conflict of interests

None to declare.

## Appendix 1: search strategies

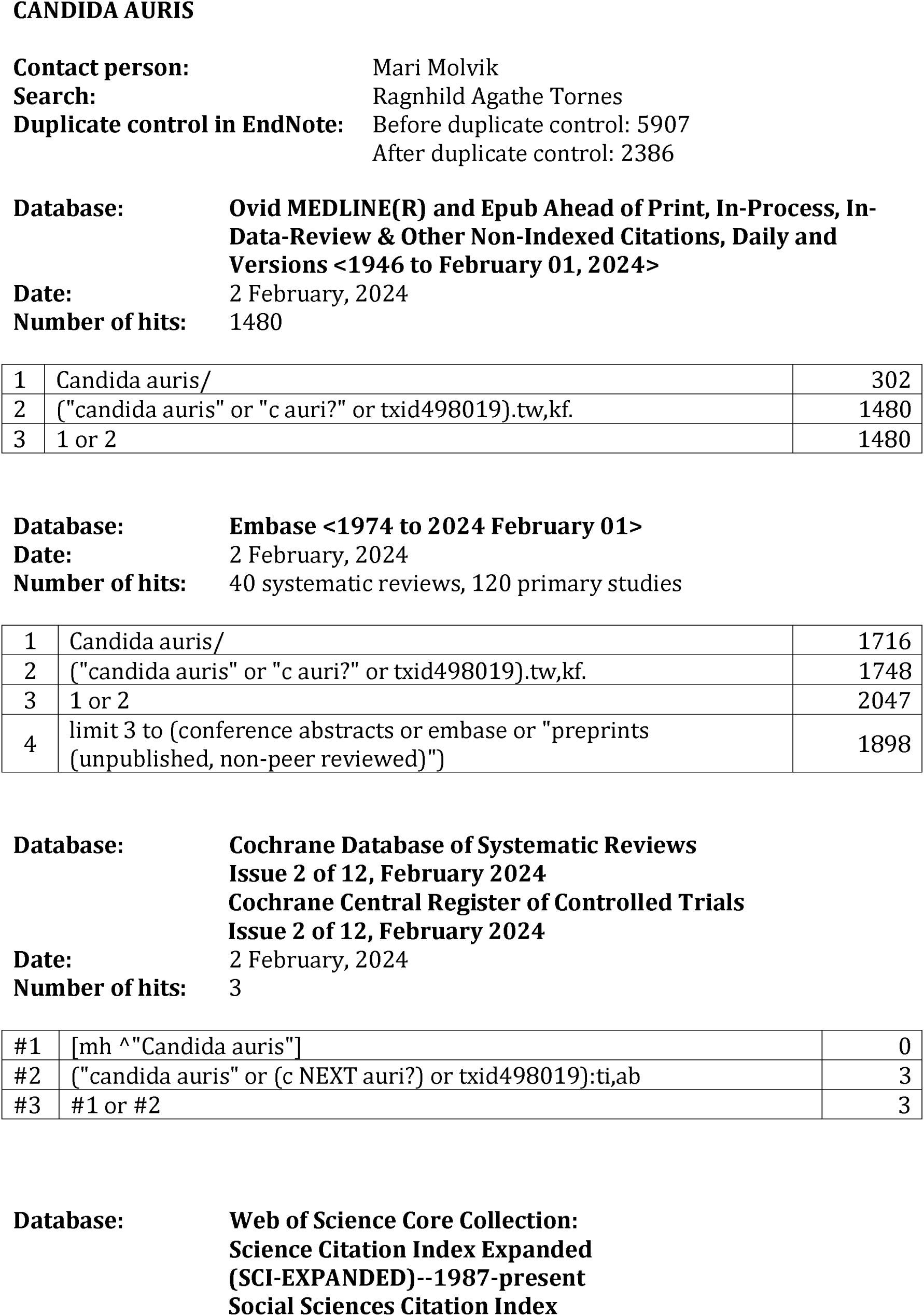
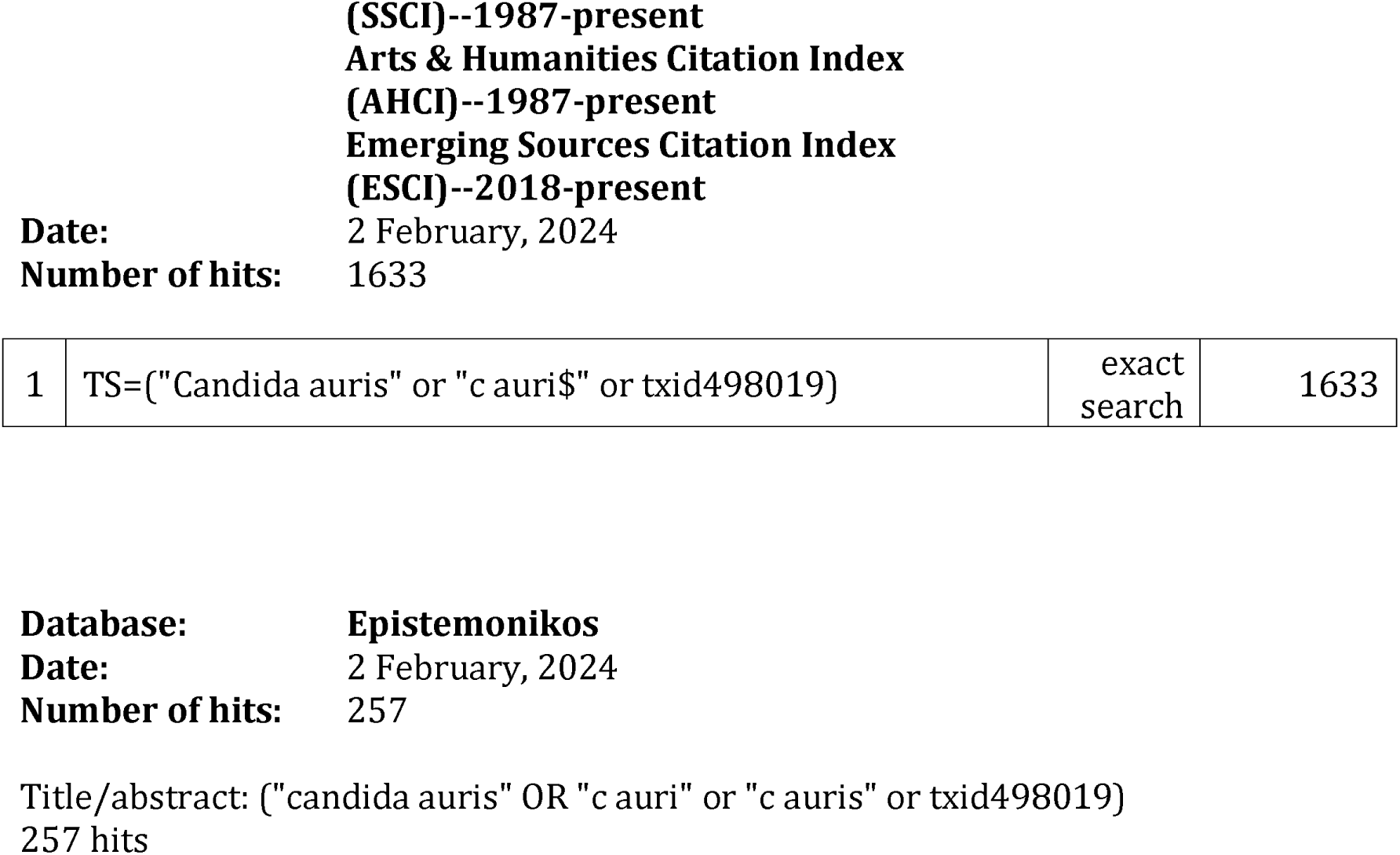

## Notes

### Competing Interest Statement

The authors have declared no competing interest.

### Funding Statement

This study was internally funded.

### Summary of Updates

Several changes in this version, but the major are: Merging results and discussion into a new section called 'Findings', and adding data extraction and discussion on diagnostic methods.

## References

1. Page MJ, McKenzie JE, Bossuyt PM, Boutron I, Hoffmann TC, Mulrow CD, et al. The PRISMA 2020 statement: an updated guideline for reporting systematic reviews. Bmj 2021;372:n71. DOI: 10.1136/bmj.n71

2. De Gaetano S, Midiri A, Mancuso G, Avola MG, Biondo C. Candida auris Outbreaks: Current Status and Future Perspectives. Microorganisms 2024;12(5). DOI: https://www.mdpi.com/2076-2607/12/5/927

3. Satoh K, Makimura K, Hasumi Y, Nishiyama Y, Uchida K, Yamaguchi H. Candida auris sp. nov., a novel ascomycetous yeast isolated from the external ear canal of an inpatient in a Japanese hospital. Microbiol Immunol 2009;53(1):41–4. DOI: 10.1111/j.1348-0421.2008.00083.x

4. Lee WG, Shin JH, Uh Y, Kang MG, Kim SH, Park KH, et al. First three reported cases of nosocomial fungemia caused by Candida auris. J Clin Microbiol 2011;49(9):3139–42. DOI: 10.1128/jcm.00319-11

5. Chowdhary A, Sharma C, Duggal S, Agarwal K, Prakash A, Singh PK, et al. New Clonal Strain of *Candida auris*, Delhi, India. Emerging Infectious Disease journal 2013;19(10):1670. DOI: 10.3201/eid1910.130393

6. Centers for Disease Control and Prevention (CDC). Antibiotic resistance threats in the United States [nettdokument]. Atlanta: U.S Department of Health and Human Services [cited 05.05.2024]. Available from: www.cdc.gov/DrugResistance/Biggest-Threats.html.

7. 7. World Health Organization (WHO). WHO fungal priority pathogens list to guide research, development and public health action [nettdokument]. Geneva[cited 05.05.2024]. Available from: https://iris.who.int/bitstream/handle/10665/363682/9789240060241-eng.pdf?sequence=1

8. Osei S. Candida auris: A systematic review and meta-analysis of current updates on an emerging multidrug-resistant pathogen. MicrobiologyOpen 2018;7(4):e00578. DOI: 10.1002/mbo3.578

9. Du H, Bing J, Hu T, Ennis CL, Nobile CJ, Huang G. Candida auris: Epidemiology, biology, antifungal resistance, and virulence. PLoS Pathog 2020;16(10):e1008921. DOI: 10.1371/journal.ppat.1008921

10. Horton MV, Nett JE. Candida auris infection and biofilm formation: going beyond the surface. Curr Clin Microbiol Rep 2020;7(3):51–6. DOI: 10.1007/s40588-020-00143-7

11. Kohlenberg A, Monnet DL, Plachouras D. Increasing number of cases and outbreaks caused by Candida auris in the EU/EEA, 2020 to 2021. Euro Surveill 2022;27(46). DOI: 10.2807/1560-7917.Es.2022.27.46.2200846

12. Suphavilai C, Ko KKK, Lim KM, Tan MG, Boonsimma P, Chu JJK, et al. Detection and characterisation of a sixth Candida auris clade in Singapore: a genomic and phenotypic study. Lancet Microbe 2024;5(9):100878. DOI: 10.1016/s2666-5247(24)00101-0

13. Jones CR, Neill C, Borman AM, Budd EL, Cummins M, Fry C, et al. The laboratory investigation, management, and infection prevention and control of Candida auris: a narrative review to inform the 2024 national guidance update in England. J Med Microbiol 2024;73(5). DOI: 10.1099/jmm.0.001820

14. Meijer EF, Voss A. Should all hospitalised patients colonised with Candida auris be considered for isolation? Eurosurveillance 2024;29(45):2400729. DOI: 10.2807/1560-7917.ES.2024.29.45.2400729

15. Desnos-Ollivier M, Fekkar A, Bretagne S. Earliest case of Candida auris infection imported in 2007 in Europe from India prior to the 2009 description in Japan. J Mycol Med 2021;31(3):101139. DOI: 10.1016/j.mycmed.2021.101139

16. Eyre DW, Sheppard AE, Madder H, Moir I, Moroney R, Quan TP, et al. A Candida auris Outbreak and Its Control in an Intensive Care Setting. New England Journal of Medicine 2018;379(14):1322–31. DOI: 10.1056/NEJMoa1714373

17. European Centre for Disease Prevention and Control (ECDC). Candida auris outbreak in healthcare facilities in northern Italy, 2019 - 2021 [nettdokument]. Stockholm[cited]. Available from: https://www.ecdc.europa.eu/sites/default/files/documents/RRA-candida-auris-Feb2022.pdf

18. The EndNote Team. EndNote. EndNote 20 ed. Philadelphia, PA: Clarivate; 2013.

19. 19. EPPI Reviewer. [cited]. Available from: https://eppi.ioe.ac.uk/cms/Default.aspx?tabid=2914

20. Vaseghi N, Sharifisooraki J, Khodadadi H, Nami S, Safari F, Ahangarkani F, et al. Global prevalence and subgroup analyses of coronavirus disease (COVID-19) associated Candida auris infections (CACa): A systematic review and meta-analysis. Mycoses 2022;65(7):683–703. DOI: 10.1111/myc.13471

21. Vinayagamoorthy K, Pentapati KC, Prakash H. Prevalence, risk factors, treatment and outcome of multidrug resistance Candida auris infections in Coronavirus disease (COVID-19) patients: A systematic review. Mycoses 2022;65(6):613–24. DOI: 10.1111/myc.13447

22. Bergeron G, Bloch D, Murray K, Kratz M, Parton H, Ackelsberg J, et al. Candida auris Colonization After Discharge to a Community Setting: New York City, 2017-2019. Open Forum Infectious Diseases 2021;8(1):ofaa620. DOI: 10.1093/ofid/ofaa620

23. Vallabhaneni S, Kallen A, Tsay S, Chow N, Welsh R, Kerins J, et al. Investigation of the First Seven Reported Cases of Candida auris, a Globally Emerging Invasive, Multidrug-Resistant Fungus - United States, May 2013-August 2016. MMWR - Morbidity & Mortality Weekly Report 2016;65(44):1234–7. DOI: 10.15585/mmwr.mm6544e1

24. Jung J, Kim MJ, Kim JY, Lee JY, Kwak SH, Hong MJ, et al. Candida auris colonization or infection of the ear: A single-center study in South Korea from 2016 to 2018. Medical Mycology 2020;58(1):124–7. DOI: 10.1093/mmy/myz020

25. Adams E, Quinn M, Tsay S, Poirot E, Chaturvedi S, Southwick K, et al. Candida auris in Healthcare Facilities, New York, USA, 2013-2017. Emerging Infectious Diseases 2018;24(10):1816-24. DOI: 10.3201/eid2410.180649

26. Arenas SP, Persad PJ, Patel S, Parekh DJ, Ferreira TBD, Farinas M, et al. Persistent colonization of Candida auris among inpatients rescreened as part of a weekly surveillance program. Infection Control & Hospital Epidemiology 2023:1–4. DOI: 10.1017/ice.2023.251

27. Pacilli M, Kerins JL, Clegg WJ, Walblay KA, Adil H, Kemble SK, et al. Regional Emergence of Candida auris in Chicago and Lessons Learned From Intensive Follow-up at 1 Ventilator-Capable Skilled Nursing Facility. Clinical Infectious Diseases 2020;71(11):e718–e25. DOI: 10.1093/cid/ciaa435

28. Yadav A, Singh A, Wang Y, Haren MHV, Singh A, de Groot T, et al. Colonisation and Transmission Dynamics of Candida auris among Chronic Respiratory Diseases Patients Hospitalised in a Chest Hospital, Delhi, India: A Comparative Analysis of Whole Genome Sequencing and Microsatellite Typing. Journal of Fungi 2021;7(2):26. DOI: 10.3390/jof7020081

29. 29. U.S Center for Disease Control and Prevention (CDC). Infection Control Guidance: Candida auris[cited]. Available from: https://www.cdc.gov/candida-auris/hcp/infection-control/index.html

30. Kenters N, Kiernan M, Chowdhary A, Denning DW, Peman J, Saris K, et al. Control of Candida auris in healthcare institutions: Outcome of an International Society for Antimicrobial Chemotherapy expert meeting. Int J Antimicrob Agents 2019;54(4):400–6. DOI: 10.1016/j.ijantimicag.2019.08.013

31. Rossow J, Ostrowsky B, Adams E, Greenko J, McDonald R, Vallabhaneni S, et al. Factors Associated With Candida auris Colonization and Transmission in Skilled Nursing Facilities With Ventilator Units, New York, 2016-2018. Clinical Infectious Diseases 2021;72(11):e753–e60. DOI: 10.1093/cid/ciaa1462

32. Pacilli M, Walblay K, Adil H, Xydis S, Kerins J, Valley A, et al. Repeated Prevalence Surveys and Admission Screening for Candida auris at One Long-Term Acute-Care Hospital, Chicago, 2016-2019. Infection Control and Hospital Epidemiology 2020;41:S14–S5. DOI: 10.1017/ice.2020.487

33. Townsend JO, Morillo A, Braithwaite LK, Boodoosingh S, Neil A, Widla J, et al. Identification of Candida auris in a foreign repatriated patient to Ontario, Canada and infection control strategies to prevent transmission. Canadian Journal of Infection Control 2021;36(4):184–7.

34. Escandon P, Chow NA, Caceres DH, Gade L, Berkow EL, Armstrong P, et al. Molecular Epidemiology of Candida auris in Colombia Reveals a Highly Related, Countrywide Colonization With Regional Patterns in Amphotericin B Resistance. Clinical Infectious Diseases 2019;68(1):15–21. DOI: 10.1093/cid/ciy411

35. Alshamrani MM, El-Saed A, Mohammed A, Alghoribi MF, Al Johani SM, Cabanalan H, et al. Management of Candida auris outbreak in a tertiary-care setting in Saudi Arabia. Infection Control & Hospital Epidemiology 2021;42(2):149–55. DOI: 10.1017/ice.2020.414

36. Eckbo EJ, Wong T, Bharat A, Cameron-Lane M, Hoang L, Dawar M, et al. First reported outbreak of the emerging pathogen Candida auris in Canada. American Journal of Infection Control 2021;49(6):804–7. DOI: 10.1016/j.ajic.2021.01.013

37. Worth LJ, Harrison SJ, Dickinson M, van Diemen A, Breen J, Harper S, et al. Candida auris in an Australian health care facility: importance of screening high risk patients. Medical Journal of Australia 2020;212(11):510–1.e1. DOI: 10.5694/mja2.50612

38. Biswal M, Rudramurthy SM, Jain N, Shamanth AS, Sharma D, Jain K, et al. Controlling a possible outbreak of Candida auris infection: lessons learnt from multiple interventions. Journal of Hospital Infection 2017;97(4):363–70. DOI: 10.1016/j.jhin.2017.09.009

39. Lee EH, Choi MH, Lee KH, Kim D, Jeong SH, Song YG, et al. Intrahospital transmission and infection control of Candida auris originating from a severely infected COVID-19 patient transferred abroad. Journal of Hospital Infection 2024;143:140–9. DOI: 10.1016/j.jhin.2023.10.017

40. Schelenz S, Hagen F, Rhodes JL, Abdolrasouli A, Chowdhary A, Hall A, et al. First hospital outbreak of the globally emerging Candida auris in a European hospital. Antimicrobial Resistance & Infection Control 2016;5:35.

41. Ruiz-Gaitan A, Moret AM, Tasias-Pitarch M, Aleixandre-Lopez AI, Martinez-Morel H, Calabuig E, et al. An outbreak due to Candida auris with prolonged colonisation and candidaemia in a tertiary care European hospital. Mycoses 2018;61(7):498–505. DOI: 10.1111/myc.12781

42. McDougal AN, DeMaet MA, Garcia B, York T, Iverson T, Ojo O, et al. A cluster investigation of Candida auris among hospitalized incarcerated patients. Antimicrobial Stewardship & Healthcare Epidemiology : ASHE 2023;3(1):e244. DOI: 10.1017/ash.2023.520

43. Aldejohann AM, Wiese-Posselt M, Gastmeier P, Kurzai O. Expert recommendations for prevention and management of Candida auris transmission. Mycoses 2022;65(6):590–8. DOI: 10.1111/myc.13445

44. 44. European Centre for Disease Prevention and Control (ECDC). Candida auris in healthcare settings – Europe[cited]. Available from: https://www.ecdc.europa.eu/sites/default/files/documents/RRA-Candida-auris-European-Union-countries.pdf

45. Centers for Disease Control and Prevention (CDC). Public Health Strategies to Prevent the Spread of Novel and Targeted Multidrug resistant Organisms (MDROs)[cited]. Available from: https://www.cdc.gov/healthcare-associated-infections/media/pdfs/Health-Response-Prevent-MDRO-508.pdf

46. Lyman M, Forsberg K, Sexton DJ, Chow NA, Lockhart SR, Jackson BR, et al. Worsening Spread of Candida auris in the United States, 2019 to 2021. Annals of Internal Medicine 2023;176(4):489–95. DOI: 10.7326/m22-3469

47. Hinrichs C, Wiese-Posselt M, Graf B, Geffers C, Weikert B, Enghard P, et al. Successful control of Candida auris transmission in a German COVID-19 intensive care unit. Mycoses 2022;65(6):643–9. DOI: 10.1111/myc.13443

48. Adam RD, Revathi G, Okinda N, Fontaine M, Shah J, Kagotho E, et al. Analysis of Candida auris fungemia at a single facility in Kenya. International Journal of Infectious Diseases 2019;85:182–7. DOI: 10.1016/j.ijid.2019.06.001

49. Allaw F, Haddad SF, Habib N, Moukarzel P, Naji NS, Kanafani ZA, et al. COVID-19 and C. auris: A Case-Control Study from a Tertiary Care Center in Lebanon. Microorganisms 2022;10(5):11. DOI: 10.3390/microorganisms10051011

50. Alvarez-Moreno CA, Morales-Lopez S, Rodriguez GJ, Rodriguez JY, Robert E, Picot C, et al. The Mortality Attributable to Candidemia in C. auris Is Higher than That in Other Candida Species: Myth or Reality? Journal of Fungi 2023;9(4):31. DOI: 10.3390/jof9040430

51. Caceres DH, Rivera SM, Armstrong PA, Escandon P, Chow NA, Ovalle MV, et al. Case-Case Comparison of Candida auris Versus Other Candida Species Bloodstream Infections: Results of an Outbreak Investigation in Colombia. Mycopathologia 2020;185(5):917–23. DOI: 10.1007/s11046-020-00478-1

52. Farooqi JQ, Soomro AS, Baig MA, Sajjad SF, Hamid K, Jabeen K, et al. Outbreak investigation of Candida auris at a tertiary care hospital in Karachi, Pakistan. Journal of Infection Prevention 2020;21(5):189–95. DOI: 10.1177/1757177420935639

53. Moin S, Farooqi J, Rattani S, Nasir N, Zaka S, Jabeen K. C. auris and non-C. auris candidemia in hospitalized adult and pediatric COVID-19 patients; single center data from Pakistan. Medical Mycology 2021;59(12):1238–42. DOI: 10.1093/mmy/myab057

54. Parak A, Stacey SL, Chibabhai V. Clinical and laboratory features of patients with Candida auris cultures, compared to other Candida, at a South African Hospital. Journal of Infection in Developing Countries 2022;16(1):213–21. DOI: 10.3855/jidc.14917

55. Ruiz-Gaitan A, Martinez H, Moret AM, Calabuig E, Tasias M, Alastruey-Izquierdo A, et al. Detection and treatment of Candida auris in an outbreak situation: risk factors for developing colonization and candidemia by this new species in critically ill patients. Expert Review of Antiinfective Therapy 2019;17(4):295–305. DOI: 10.1080/14787210.2019.1592675

56. Sayeed MA, Farooqi J, Jabeen K, Mahmood SF. Comparison of risk factors and outcomes of Candida auris candidemia with non-Candida auris candidemia: A retrospective study from Pakistan. Medical Mycology 2020;58(6):721–9. DOI: 10.1093/mmy/myz112

57. Shastri PS, Shankarnarayan SA, Oberoi J, Rudramurthy SM, Wattal C, Chakrabarti A. Candida auris candidaemia in an intensive care unit - Prospective observational study to evaluate epidemiology, risk factors, and outcome. Journal of Critical Care 2020;57:42–8. DOI: 10.1016/j.jcrc.2020.01.004

58. Simon SP, Li R, Silver M, Andrade J, Tharian B, Fu L, et al. Comparative Outcomes of Candida auris Bloodstream Infections: A Multicenter Retrospective Case-Control Study. Clinical Infectious Diseases 2023;76(3):e1436–e43. DOI: 10.1093/cid/ciac735

59. Taori SK, Khonyongwa K, Hayden I, Athukorala GDA, Letters A, Fife A, et al. Candida auris outbreak: Mortality, interventions and cost of sustaining control. Journal of Infection 2019;79(6):601–11. DOI: 10.1016/j.jinf.2019.09.007

60. van Schalkwyk E, Mpembe RS, Thomas J, Shuping L, Ismail H, Lowman W, et al. Epidemiologic Shift in Candidemia Driven by Candida auris, South Africa, 2016-2017^1^. Emerging Infectious Diseases 2019;25(9):1698–707. DOI: 10.3201/eid2509.190040

61. Bender R, Lange S. Adjusting for multiple testing—when and how? Journal of clinical epidemiology 2001;54(4):343–9.

62. Westreich D, Greenland S. The table 2 fallacy: presenting and interpreting confounder and modifier coefficients. American journal of epidemiology 2013;177(4):292–8.

63. Leonhard SE, Chong GL, Foudraine DE, Bode LGM, Croughs P, Popping S, et al. Proposal for a screening protocol for Candida auris colonization. Journal of Hospital Infection 2024;27:27. DOI: 10.1016/j.jhin.2023.12.019

64. 64. Norwegian Institute of Public Health (NIPH). Anbefalt screening for resistente mikrober hos pasienter overflyttet fra utlandet[cited]. Available from: https://www.fhi.no/sm/smittevern-i-helsetjenesten/anbefalt-screening-for-resistente-mikrober-hos-pasienter-overflyttet-fra-ut/

65. Hamprecht A, Barber AE, Mellinghoff SC, Thelen P, Walther G, Yu Y, et al. Candida auris in Germany and Previous Exposure to Foreign Healthcare. Emerging Infectious Diseases 2019;25(9):1763–5. DOI: 10.3201/eid2509.190262

66. Turbett ISE, Becker DSM, Belford MTB, Kelly RTM, Desrosiers MTL, Oliver RNE, et al. Evaluation of Candida auris acquisition in US international travellers using a culture-based screening protocol1. Journal of Travel Medicine 2022;29(1):17. DOI: 10.1093/jtm/taab186

67. Yadav A, Jain K, Wang Y, Pawar K, Kaur H, Sharma KK, et al. Candida auris on Apples: Diversity and Clinical Significance. mBio 2022;13(2):e0051822. DOI: 10.1128/mbio.00518-22

68. 68. UK Health Security Agency (UKHSA). Candida auris: a review of recent literature[cited]. Available from: https://www.gov.uk/government/consultations/candida-auris-update-to-management-guidance/candida-auris-a-review-of-recent-literature

69. Chakrabarti A, Singh S. Multidrug-resistant Candida auris: an epidemiological review. Expert Rev Anti Infect Ther 2020;18(6):551–62. DOI: 10.1080/14787210.2020.1750368

70. Armstrong PA, Rivera SM, Escandon P, Caceres DH, Chow N, Stuckey MJ, et al. Hospital-Associated Multicenter Outbreak of Emerging Fungus Candida auris, Colombia, 2016. Emerging Infectious Diseases 2019;25(7):07. DOI: 10.3201/eid2507.180491

71. de Melo CC, de Sousa BR, da Costa GL, Oliveira MME, de Lima-Neto RG. Colonized patients by Candida auris: Third and largest outbreak in Brazil and impact of biofilm formation. Frontiers in Cellular & Infection Microbiology 2023;13:1033707. DOI: 10.3389/fcimb.2023.1033707

72. Goulart MA, El Itani R, Buchanan SR, Brown DS, Hays AK, King WB, et al. Identification and infection control response to Candida auris at an academic level I trauma center. American Journal of Infection Control 2023;28:28. DOI: 10.1016/j.ajic.2023.11.017

73. McPherson TD, Walblay KA, Roop E, Soglin D, Valley A, Logan LK, et al. Notes from the Field: Candida auris and Carbapenemase-Producing Organism Prevalence in a Pediatric Hospital Providing Long-Term Transitional Care - Chicago, Illinois, 2019. MMWR - Morbidity & Mortality Weekly Report 2020;69(34):1180–1. DOI: 10.15585/mmwr.mm6934a5

74. Southwick K, Ostrowsky B, Greenko J, Adams E, Lutterloh E, Denis RJ, et al. A description of the first Candida auris-colonized individuals in New York State, 2016-2017. American Journal of Infection Control 2022;50(3):358–60. DOI: 10.1016/j.ajic.2021.10.037

75. Rowlands J, Dufort E, Chaturvedi S, Zhu Y, Quinn M, Bucher C, et al. Candida auris admission screening pilot in select units of New York City health care facilities, 2017-2019. American Journal of Infection Control 2023;51(8):866–70. DOI: 10.1016/j.ajic.2023.01.012

76. Zhu Y, O’Brien B, Leach L, Clarke A, Bates M, Adams E, et al. Laboratory Analysis of an Outbreak of Candida auris in New York from 2016 to 2018: Impact and Lessons Learned. Journal of Clinical Microbiology 2020;58(4):25. DOI: 10.1128/jcm.01503-19

77. Proctor DM, Dangana T, Sexton DJ, Fukuda C, Yelin RD, Stanley M, et al. Integrated genomic, epidemiologic investigation of Candida auris skin colonization in a skilled nursing facility. Nature Medicine 2021;27(8):1401–9. DOI: 10.1038/s41591-021-01383-w

78. Piatti G, Sartini M, Cusato C, Schito AM. Colonization by Candida auris in critically ill patients: role of cutaneous and rectal localization during an outbreak. Journal of Hospital Infection 2022;120:85–9. DOI: 10.1016/j.jhin.2021.11.004

79. Alshahrani FS, Elgujja AA, Alsubaie S, Ezreqat SA, Albarraq AM, Barry M, et al. Description of Candida auris Occurrence in a Tertiary Health Institution in Riyadh, Saudi Arabia. Healthcare 2023;11(24):12. DOI: 10.3390/healthcare11243150

80. 80. Centers for Disease Control and Prevention (CDC). Screening for Colonization in Healthcare Facilities[cited]. Available from: https://www.cdc.gov/candida-auris/hcp/screening-hcp/index.html#cdc_generic_section_2-screening

81. 81. Pan American Health Organization (PAHO) / World Health Organization (WHO). Candida auris outbreaks in health care services in the context of the COVID-19 pandemic[cited]. Available from: https://iris.paho.org/bitstream/handle/10665.2/53377/EpiUpdate6February2021_eng.pdf?sequence=1&isAllowed=y

82. Sansom SE, Gussin GM, Schoeny M, Singh RD, Adil H, Bell P, et al. Rapid Environmental Contamination with Candida auris and Multidrug-Resistant Bacterial Pathogens Near Colonized Patients. Clinical Infectious Diseases 2023;06:06. DOI: 10.1093/cid/ciad752

83. Ahmad S, Khan Z, Al-Sweih N, Alfouzan W, Joseph L. Candida auris in various hospitals across Kuwait and their susceptibility and molecular basis of resistance to antifungal drugs. Mycoses 2020;63(1):104–12. DOI: 10.1111/myc.13022

84. Al Maani A, Paul H, Al-Rashdi A, Wahaibi AA, Al-Jardani A, Al Abri AMA, et al. Ongoing Challenges with Healthcare-Associated Candida auris Outbreaks in Oman. Journal of Fungi 2019;5(4):23. DOI: 10.3390/jof5040101

85. Alfouzan W, Ahmad S, Dhar R, Asadzadeh M, Almerdasi N, Abdo NM, et al. Molecular Epidemiology of Candida Auris Outbreak in a Major Secondary-Care Hospital in Kuwait. Journal of Fungi 2020;6(4):21. DOI: 10.3390/jof6040307

86. Allaw F, Kara Z, Ibrahim A, Tannous J, Taleb H, Bizri AR, et al. First Candida auris Outbreak during a COVID-19 Pandemic in a Tertiary-Care Center in Lebanon. Pathogens 2021;10(2):03. DOI: 10.3390/pathogens10020157

87. Almaghrabi RS, Albalawi R, Mutabagani M, Atienza E, Aljumaah S, Gade L, et al. Molecular characterisation and clinical outcomes of Candida auris infection: Single-centre experience in Saudi Arabia. Mycoses 2020;63(5):452–60. DOI: 10.1111/myc.13065

88. Al-Rashdi A, Al-Maani A, Al-Wahaibi A, Alqayoudhi A, Al-Jardani A, Al-Abri S. Characteristics, Risk Factors, and Survival Analysis of Candida auris Cases: Results of One-Year National Surveillance Data from Oman. Journal of Fungi 2021;7(1):07. DOI: 10.3390/jof7010031

89. Alvarado-Socarras JL, Vargas-Soler JA, Franco-Paredes C, Villegas-Lamus KC, Rojas-Torres JP, Rodriguez-Morales AJ. A Cluster of Neonatal Infections Caused by Candida auris at a Large Referral Center in Colombia. Journal of the Pediatric Infectious Diseases Societ 2021;10(5):549–55. DOI: 10.1093/jpids/piaa152

90. Amer HA, AlFaraj S, Alboqami K, Alshakarh F, Alsalam M, Kumar D, et al. Characteristics and Mitigation Measures of Candida auris Infection: Descriptive Analysis from a Quaternary Care Hospital in Saudi Arabia, 2021-2022. Journal of Epidemiology and Global Health 2023;13(4):825–30. DOI: 10.1007/s44197-023-00154-9

91. Arauz AB, Caceres DH, Santiago E, Armstrong P, Arosemena S, Ramos C, et al. Isolation of Candida auris from 9 patients in Central America: Importance of accurate diagnosis and susceptibility testing. Mycoses 2018;61(1):44–7. DOI: 10.1111/myc.12709

92. Arensman K, Miller JL, Chiang A, Mai N, Levato J, LaChance E, et al. Clinical Outcomes of Patients Treated for Candida auris Infections in a Multisite Health System, Illinois, USA. Emerging Infectious Diseases 2020;26(5):866–71. DOI: 10.3201/eid2605.191588

93. Asadzadeh M, Mokaddas E, Ahmad S, Abdullah AA, de Groot T, Meis JF, et al. Molecular characterisation of Candida auris isolates from immunocompromised patients in a tertiary-care hospital in Kuwait reveals a novel mutation in FKS1 conferring reduced susceptibility to echinocandins. Mycoses 2022;65(3):331–43. DOI: 10.1111/myc.13419

94. Barantsevich NE, Orlova OE, Shlyakhto EV, Johnson EM, Woodford N, Lass-Floerl C, et al. Emergence of Candida auris in Russia. Journal of Hospital Infection 2019;102(4):445–8. DOI: 10.1016/j.jhin.2019.02.021

95. Barantsevich NE, Vetokhina AV, Ayushinova NI, Orlova OE, Barantsevich EP. Candida auris Bloodstream Infections in Russia. Antibiotics 2020;9(9):30. DOI: 10.3390/antibiotics9090557

96. Benedict K, Forsberg K, Gold JAW, Baggs J, Lyman M. Candida auris-Associated Hospitalizations, United States, 2017-2022. Emerging Infectious Diseases 2023;29(7):1485-7. DOI: 10.3201/eid2907.230540

97. Berrio I, Caceres DH, Coronell RW, Salcedo S, Mora L, Marin A, et al. Bloodstream Infections With Candida auris Among Children in Colombia: Clinical Characteristics and Outcomes of 34 Cases. Journal of the Pediatric Infectious Diseases Societ 2021;10(2):151–4. DOI: 10.1093/jpids/piaa038

98. Bing J, Du H, Guo P, Hu T, Xiao M, Lu S, et al. Candida auris-associated hospitalizations and outbreaks, China, 2018-2023. Emerging Microbes & Infections 2024;13(1):2302843. DOI: 10.1080/22221751.2024.2302843

99. Biran R, Cohen R, Finn T, Brosh-Nissimov T, Rahav G, Yahav D, et al. Nationwide Outbreak of Candida auris Infections Driven by COVID-19 Hospitalizations, Israel, 2021-2022. Emerging Infectious Diseases 2023;29(7):1297–301. DOI: 10.3201/eid2907.221888

100. Briano F, Magnasco L, Sepulcri C, Dettori S, Dentone C, Mikulska M, et al. Candida auris Candidemia in Critically Ill, Colonized Patients: Cumulative Incidence and Risk Factors. Infectious Diseases & Therapy 2022;11(3):1149–60. DOI: 10.1007/s40121-022-00625-9

101. Calvo B, Melo AS, Perozo-Mena A, Hernandez M, Francisco EC, Hagen F, et al. First report of Candida auris in America: Clinical and microbiological aspects of 18 episodes of candidemia. Journal of Infection 2016;73(4):369–74. DOI: 10.1016/j.jinf.2016.07.008

102. Chakrabarti A, Sood P, Rudramurthy SM, Chen S, Jillwin J, Iyer R, et al. Characteristics, outcome and risk factors for mortality of paediatric patients with ICU-acquired candidemia in India: A multicentre prospective study. Mycoses 2020;63(11):1149–63. DOI: 10.1111/myc.13145

103. Chandramati J, Sadanandan L, Kumar A, Ponthenkandath S. Neonatal Candida auris infection: Management and prevention strategies - A single centre experience. Journal of Paediatrics & Child Health 2020;56(10):1565–9. DOI: 10.1111/jpc.15019

104. Chibabhai V. Incidence of candidemia and prevalence of azole-resistant candidemia at a tertiary South African hospital - A retrospective laboratory analysis 2016-2020. Southern African Journal of Infectious Diseases 2022;37(1):326. DOI: 10.4102/sajid.v37i1.326

105. Chow NA, Gade L, Tsay SV, Forsberg K, Greenko JA, Southwick KL, et al. Multiple introductions and subsequent transmission of multidrug-resistant Candida auris in the USA: a molecular epidemiological survey. The Lancet Infectious Diseases 2018;18(12):1377–84. DOI: 10.1016/s1473-3099(18)30597-8

106. Chowdhary A, Tarai B, Singh A, Sharma A. Multidrug-Resistant Candida auris Infections in Critically Ill Coronavirus Disease Patients, India, April-July 2020. Emerging Infectious Diseases 2020;26(11):2694–6. DOI: 10.3201/eid2611.203504

107. Corcione S, Montrucchio G, Shbaklo N, De Benedetto I, Sales G, Cedrone M, et al. First Cases of Candida auris in a Referral Intensive Care Unit in Piedmont Region, Italy. Microorganisms 2022;10(8):27. DOI: 10.3390/microorganisms10081521

108. de St Maurice A, Parti U, Anikst VE, Harper T, Mirasol R, Dayo AJ, et al. Clinical, microbiological, and genomic characteristics of clade-III Candida auris colonization and infection in southern California, 2019-2022. Infection Control & Hospital Epidemiology 2023;44(7):1093–101. DOI: 10.1017/ice.2022.204

109. Di Pilato V, Codda G, Ball L, Giacobbe DR, Willison E, Mikulska M, et al. Molecular Epidemiological Investigation of a Nosocomial Cluster of C. auris: Evidence of Recent Emergence in Italy and Ease of Transmission during the COVID-19 Pandemic. Journal of Fungi 2021;7(2):15. DOI: 10.3390/jof7020140

110. Escandon P, Caceres DH, Espinosa-Bode A, Rivera S, Armstrong P, Vallabhaneni S, et al. Notes from the Field: Surveillance for Candida auris - Colombia, September 2016-May 2017. MMWR - Morbidity & Mortality Weekly Report 2018;67(15):459–60. DOI: 10.15585/mmwr.mm6715a6

111. Escandon P, Caceres DH, Lizarazo D, Lockhart SR, Lyman M, Duarte C. Laboratory-based surveillance of Candida auris in Colombia, 2016-2020. Mycoses 2022;65(2):222–5. DOI: 10.1111/myc.13390

112. Garcia-Bustos V, Salavert M, Ruiz-Gaitan AC, Cabanero-Navalon MD, Sigona-Giangreco IA, Peman J. A clinical predictive model of candidaemia by Candida auris in previously colonized critically ill patients. Clinical Microbiology & Infection 2020;26(11):1507–13. DOI: 10.1016/j.cmi.2020.02.001

113. Garcia-Jeldes HF, Mitchell R, McGeer A, Rudnick W, Amaratunga K, Vallabhaneni S, et al. Prevalence of Candida auris in Canadian acute care hospitals among at-risk patients, 2018. Antimicrobial Resistance & Infection Control 2020;9(1):82. DOI: 10.1186/s13756-020-00752- 3

114. Gómez CF, Albamonte PS, Navarro CD, García CS, Palop NT, Ibáñez JAA. Analysis of Candida auris candidemia cases in an Intensive Care Unit of a tertiary hospital. Revista Espanola De Anestesiologia Y Reanimacion 2021;68(8):431–6. DOI: 10.1016/j.redar.2020.10.013

115. Govender NP, Magobo RE, Mpembe R, Mhlanga M, Matlapeng P, Corcoran C, et al. Candida auris in South Africa, 2012-2016. Emerging Infectious Diseases 2018;24(11):2036–40. DOI: 10.3201/eid2411.180368

116. Hanson BM, Dinh AQ, Tran TT, Arenas S, Pronty D, Gershengorn HB, et al. Candida auris Invasive Infections during a COVID-19 Case Surge. Antimicrobial Agents & Chemotherapy 2021;65(10):e0114621. DOI: 10.1128/aac.01146-21

117. Kaki R. Risk factors and mortality of the newly emerging Candida auris in a university hospital in Saudi Arabia. Mycology 2023;14(3):256–63. DOI: 10.1080/21501203.2023.2227218

118. Kekana D, Naicker SD, Shuping L, Velaphi S, Nakwa FL, Wadula J, et al. Candida auris Clinical Isolates Associated with Outbreak in Neonatal Unit of Tertiary Academic Hospital, South Africa. Emerging Infectious Diseases 2023;29(10):2044–53. DOI: 10.3201/eid2910.230181

119. Khan Z, Ahmad S, Benwan K, Purohit P, Al-Obaid I, Bafna R, et al. Invasive Candida auris infections in Kuwait hospitals: epidemiology, antifungal treatment and outcome. Infection 2018;46(5):641–50. DOI: 10.1007/s15010-018-1164-y

120. Koleri J, Petkar HM, Rahman SASHA, Rahman SAMA. Candida auris Blood stream infection- a descriptive study from Qatar. BMC Infectious Diseases 2023;23(1):513. DOI: 10.1186/s12879-023-08477-5

121. Lockhart SR, Etienne KA, Vallabhaneni S, Farooqi J, Chowdhary A, Govender NP, et al. Simultaneous Emergence of Multidrug-Resistant Candida auris on 3 Continents Confirmed by Whole-Genome Sequencing and Epidemiological Analyses. Clinical Infectious Diseases 2017;64(2):134–40. DOI: 10.1093/cid/ciw691

122. Magnasco L, Mikulska M, Giacobbe DR, Taramasso L, Vena A, Dentone C, et al. Spread of Carbapenem-Resistant Gram-Negatives and Candida auris during the COVID-19 Pandemic in Critically Ill Patients: One Step Back in Antimicrobial Stewardship? Microorganisms 2021;9(1):03. DOI: 10.3390/microorganisms9010095

123. Magnasco L, Mikulska M, Sepulcri C, Ullah N, Giacobbe DR, Vena A, et al. Frequency of Detection of Candida auris Colonization Outside a Highly Endemic Setting: What Is the Optimal Strategy for Screening of Carriage? Journal of Fungi 2023;10(1):29. DOI: 10.3390/jof10010026

124. Magobo R, Mhlanga M, Corcoran C, Govender NP. Multilocus sequence typing of azole-resistant Candida auris strains, South Africa. Southern African Journal of Infectious Diseases 2020;35(1):116. DOI: 10.4102/sajid.v35i1.116

125. Mohsin J, Weerakoon S, Ahmed S, Puts Y, Al Balushi Z, Meis JF, et al. A Cluster of Candida auris Blood Stream Infections in a Tertiary Care Hospital in Oman from 2016 to 2019. Antibiotics 2020;9(10):24. DOI: 10.3390/antibiotics9100638

126. Morales-Lopez SE, Parra-Giraldo CM, Ceballos-Garzon A, Martinez HP, Rodriguez GJ, Alvarez-Moreno CA, et al. Invasive Infections with Multidrug-Resistant Yeast Candida auris, Colombia. Emerging Infectious Diseases 2017;23(1):162–4. DOI: 10.3201/eid2301.161497

127. Mulet B, J V, Tormo P, Salvador G, Guna S, M DR, et al. Candida auris from colonisation to candidemia: A four-year study. Mycoses 2023;66(10):882–90. DOI: 10.1111/myc.13626

128. Mulet B, J V, Tormo P, Salvador G, Herrero R, Abril Lopez de Medrano V, et al. Characteristics and Management of Candidaemia Episodes in an Established Candida auris Outbreak. Antibiotics 2020;9(9):30. DOI: 10.3390/antibiotics9090558

129. Munshi A, Almadani F, Ossenkopp J, Alharbi M, Althaqafi A, Alsaedi A, et al. Risk factors, antifungal susceptibility, complications, and outcome of Candida auris bloodstream infection in a tertiary care center in the western region of Saudi Arabia. Journal of Infection and Public Health 2024;17(1):182–8. DOI: 10.1016/j.jiph.2023.11.021

130. Nobrega de Almeida J, Jr, Brandao IB, Francisco EC, de Almeida SLR, de Oliveira Dias P, et al. Axillary Digital Thermometers uplifted a multidrug-susceptible Candida auris outbreak among COVID-19 patients in Brazil. Mycoses 2021;64(9):1062–72. DOI: 10.1111/myc.13320

131. Ortiz-Roa C, Valderrama-Rios MC, Sierra-Umana SF, Rodriguez JY, Muneton-Lopez GA, Solorzano-Ramos CA, et al. Mortality Caused by Candida auris Bloodstream Infections in Comparison with Other Candida Species, a Multicentre Retrospective Cohort. Journal of Fungi 2023;9(7):29. DOI: 10.3390/jof9070715

132. Pandya N, Cag Y, Pandak N, Pekok AU, Poojary A, Ayoade F, et al. International Multicentre Study of Candida auris Infections. Journal of Fungi 2021;7(10):19. DOI: 10.3390/jof7100878

133. Park JY, Bradley N, Brooks S, Burney S, Wassner C. Management of Patients with Candida auris Fungemia at Community Hospital, Brooklyn, New York, USA, 2016-2018^1^. Emerging Infectious Diseases 2019;25(3):601–2. DOI: 10.3201/eid2503.180927

134. Peng Y, Liu Y, Yu X, Fang J, Guo Z, Liao K, et al. First report of Candida auris in Guangdong, China: clinical and microbiological characteristics of 7 episodes of candidemia. Emerging Microbes & Infections 2024;13(1):2300525. DOI: 10.1080/22221751.2023.2300525

135. Prestel C, Anderson E, Forsberg K, Lyman M, de Perio MA, Kuhar D, et al. Candida auris Outbreak in a COVID-19 Specialty Care Unit - Florida, July-August 2020. MMWR - Morbidity & Mortality Weekly Report 2021;70(2):56–7. DOI: 10.15585/mmwr.mm7002e3

136. Sathyapalan DT, Antony R, Nampoothiri V, Kumar A, Shashindran N, James J, et al. Evaluating the measures taken to contain a Candida auris outbreak in a tertiary care hospital in South India: an outbreak investigational study. BMC Infectious Diseases 2021;21(1):425. DOI: 10.1186/s12879-021-06131-6

137. Sayeed MA, Farooqi J, Jabeen K, Awan S, Mahmood SF. Clinical spectrum and factors impacting outcome of Candida auris: a single center study from Pakistan. BMC Infectious Diseases 2019;19(1):384. DOI: 10.1186/s12879-019-3999-y

138. Sharp A, Muller-Pebody B, Charlett A, Patel B, Gorton R, Lambourne J, et al. Screening for Candida auris in patients admitted to eight intensive care units in England, 2017 to 2018. Euro Surveillance: Bulletin Europeen sur les Maladies Transmissibles = European Communicable Disease Bulletin 2021;26(8). DOI: 10.2807/1560-7917.Es.2021.26.8.1900730

139. Shaukat A, Al Ansari N, Al Wali W, Karic E, El Madhoun I, Mitwally H, et al. Experience of treating Candida auris cases at a general hospital in the state of Qatar. IDCases 2021;23:e01007. DOI: 10.1016/j.idcr.2020.e01007

140. Stanciu AM, Florea D, Surleac M, Paraschiv S, Otelea D, Talapan D, et al. First report of Candida auris in Romania: clinical and molecular aspects. Antimicrobial Resistance & Infection Control 2023;12(1):91. DOI: 10.1186/s13756-023-01297-x

141. Sticchi C, Raso R, Ferrara L, Vecchi E, Ferrero L, Filippi D, et al. Increasing Number of Cases Due to Candida auris in North Italy, July 2019-December 2022. Journal of Clinical Medicine 2023;12(5):28. DOI: 10.3390/jcm12051912

142. Taori SK, Rhodes J, Khonyongwa K, Szendroi A, Smith M, Borman AM, et al. First experience of implementing Candida auris real-time PCR for surveillance in the UK: detection of multiple introductions with two international clades and improved patient outcomes. Journal of Hospital Infection 2022;127:111–20. DOI: 10.1016/j.jhin.2022.06.009

143. Thomsen J, Abdulrazzaq NM, Oulhaj A, Nyasulu PS, Alatoom A, Denning DW, et al. Emergence of highly resistant Candida auris in the United Arab Emirates: a retrospective analysis of evolving national trends. Frontiers in Public Health 2023;11:1244358. DOI: 10.3389/fpubh.2023.1244358

144. Tian S, Rong C, Nian H, Li F, Chu Y, Cheng S, et al. First cases and risk factors of super yeast Candida auris infection or colonization from Shenyang, China. Emerging Microbes & Infections 2018;7(1):128. DOI: 10.1038/s41426-018-0131-0

145. Tsay S, Welsh RM, Adams EH, Chow NA, Gade L, Berkow EL, et al. Notes from the Field: Ongoing Transmission of Candida auris in Health Care Facilities - United States, June 2016- May 2017. MMWR - Morbidity & Mortality Weekly Report 2017;66(19):514–5. DOI: 10.15585/mmwr.mm6619a7

146. Umamaheshwari S, Neelambike SM, Shankarnarayan SA, Kumarswamy KS, Gopal S, Prakash H, et al. Clinical profile, antifungal susceptibility, and molecular characterization of Candida auris isolated from patients in a South Indian surgical ICU. Journal de Mycologie Medicale 2021;31(4):101176. DOI: 10.1016/j.mycmed.2021.101176

147. Villanueva-Lozano H, Trevino-Rangel RJ, Gonzalez GM, Ramirez-Elizondo MT, Lara-Medrano R, Aleman-Bocanegra MC, et al. Outbreak of Candida auris infection in a COVID-19 hospital in Mexico. Clinical Microbiology & Infection 2021;08:08. DOI: 10.1016/j.cmi.2020.12.030

148. Vu CA, Jimenez A, Anjan S, Abbo LM. Challenges and opportunities in stewardship among solid organ transplant recipients with Candida auris bloodstream infections. Transplant Infectious Disease 2022;24(5):e13919. DOI: 10.1111/tid.13919

149. Walits E, Schaefer S. Outcome of Candida auris contact investigations conducted in a 6 month period at a New York City hospital. American Journal of Infection Control 2023;12:12. DOI: 10.1016/j.ajic.2023.10.005

150. Waters A, Chommanard C, Baltozer S, Angel LC, Abdelfattah R, Lyman M, et al. Investigation of a Candida auris outbreak in a skilled nursing facility - Virginia, United States, October 2020- June 2021. American Journal of Infection Control 2023;51(4):472–4. DOI: 10.1016/j.ajic.2022.12.003

151. Zerrouki H, Ibrahim A, Rebiahi SA, Elhabiri Y, Benhaddouche DE, de Groot T, et al. Emergence of Candida auris in intensive care units in Algeria. Mycoses 2022;65(7):753–9. DOI: 10.1111/myc.13470

